# A runtime alterable epidemic model with genetic drift, waning immunity, and vaccinations

**DOI:** 10.1101/2021.06.07.21258504

**Authors:** Wayne M. Getz, Richard Salter, Ludovica Luisa Vissat, James S. Koopman, Carl P. Simon

## Abstract

In this paper, we present methods for building a Java Runtime-Alterable-Model Platform (RAMP) of complex dynamical systems. We illustrate our methods by building a multivariant SEIR (epidemic) RAMP. Underlying our RAMP is an individual-based model that includes adaptive contact rates, pathogen genetic drift, waning and cross immunity. Besides allowing parameter values, process descriptions, and scriptable runtime drivers to be easily modified during simulations, our RAMP is easily integrated into other computational platforms, such as our illustrated example with R-Studio. Processes descriptions that can be runtime altered within our SEIR RAMP include pathogen variant-dependent host shedding, environmental persistence, host transmission, and within-host pathogen mutation and replication. They also include adaptive social distancing and adaptive application of vaccination rates and variant-valency of vaccines. We present simulation results using parameter values and process descriptions relevant to the current COVID-19 pandemic. Our results suggest that if waning immunity outpaces vaccination rates, then vaccination rollouts may fail to contain the most transmissible variants, particularly if vaccine valencies do not adapt to escape mutations. Our SEIR RAMP is designed for easy-use by individuals and groups involved in formulating social-distancing and adaptive vaccination rollout policies. More generally, our RAMP concept facilitates construction of highly flexible complex systems models of all types, which can then be easily shared among researchers and policymakers as stand alone applications programs.

## 1 Introduction

Kermack and McKendrick pioneered the application of differential equations to modeling the dynamics of disease systems that included susceptible (S), infected/infectious (E/I) and recovered (R, we use V to include vaccinated) classes of individuals [1]. Subsequent extensions of their formulation include, *inter alia*, additional disease and demographic classes [2], multihost and pathogen strain considerations [3,4], spatial heterogeneity [5,6], network [7] and individual-based formulations [8, 9]. Along with these extensions has come the challenge of “not being able to see the forest for the trees” when questions beyond those pertaining to the profiles of epidemics on homogeneous, well-mixed, large populations arise. As with the current COVID-19 pandemic, these questions may relate to the emergence of new pathogen variants [10], the effects of waning and cross-immunity in hosts with different exposure histories to these variants [11], differential transmission and virulence of these variants, issues of spatial heterogeneity and host heterogeneity related to age, gender, and health status factors [12].

We only have the capacity from both technology and human comprehension points of view to understand at any one time how a limited number of factors may explain or affect epidemiological outcomes when measures are applied to mitigate the severity of disease outbreaks. Thus, we are brought to consider the issue of how to craft a model so that it has the “appropriate level of complexity” to address the questions at hand [13, 14]. We otherwise follow Einstein’s dictum that “models should be as simple as possible, but no simpler.”

To facilitate the processes of both “incorporating complexity into” and “stripping complexity out of” models, we have developed the concept of a Runtime Alterable Model Platform (RAMP). This allow us to focus on outcomes rather than on the logistics of modifying and coding models and carrying out comparative analyses. Our RAMP includes panels, windows and sliders that allow users to specify and manipulate model parameter values, modify process function descriptions, and scripting drivers for implementing sets of simulations. Further, modifications can be made both at the start of and during the course of a simulation, while protecting the integrity of the underlying code. In addition, our RAMP automates documentation of all parameter values, process descriptions, changes and actions (modifications and substitutions during simulation) in a file that is then saved at the end of each simulation. This file is then ready for later comparative analyses across sets of simulations, or within data processing environment that incorporate our RAMP as a component package, such as R-Studio.

RAMPs can be developed for models that address classes of problems, formulated using a Goldilocks principle. Thus, these classes should not be too general so that comparisons within each class require extensive alterations to models (members of the class should share significant structural properties with regard to process dynamics), but also not too specialized so that comparisons across members of the class are too limited to provide answers to question of interest. Thus we might develop different RAMPs to study genetic, morphogentics, epidemiological, evolutionary, geological, and environmental processes.

Here we provide an example of a RAMP that has sufficient breadth to investigate an array of questions pertaining to multivariant epidemiological dynamics for directly transmissible diseases, such as the current SARS-CoV-2 pandemic [15, 16], influenza [17], or Ebola [13, 18]. For simplicity, we refer to this as our M-SEIR (multivariant susceptible-exposed-infected-recovered) RAMP. For the study of water-borne or vector-borne diseases, similar but somewhat more complicated RAMPs will need to be developed. Our M-SEIR RAMP is designed to be used by individuals either with no coding skills, or with minimal coding skills if they desire to modify some of the process descriptions incorporated into the supplied platform. It is sufficiently detailed, however, to allow the user to incorporate either supplied or user-altered versions of the following processes: i) pathogen variant-specific shedding [19], environmental persistence [20], within-host replication [21] and mortality rates [22]; ii) immunological waning with variant cross immunity [23, 24]; iii) pathogen variant drift during transmission and within-host replication [25]; iv) an adaptive contact rate [26]; v.) a time-dependent, uni- or multivalent vaccine rollout [27, 28] (Fig. 1; for mathematical details see Materials and Methods, as well as our Supplementary Online File— here forth referred to as SOF—Appendix A).

**Figure 1:**
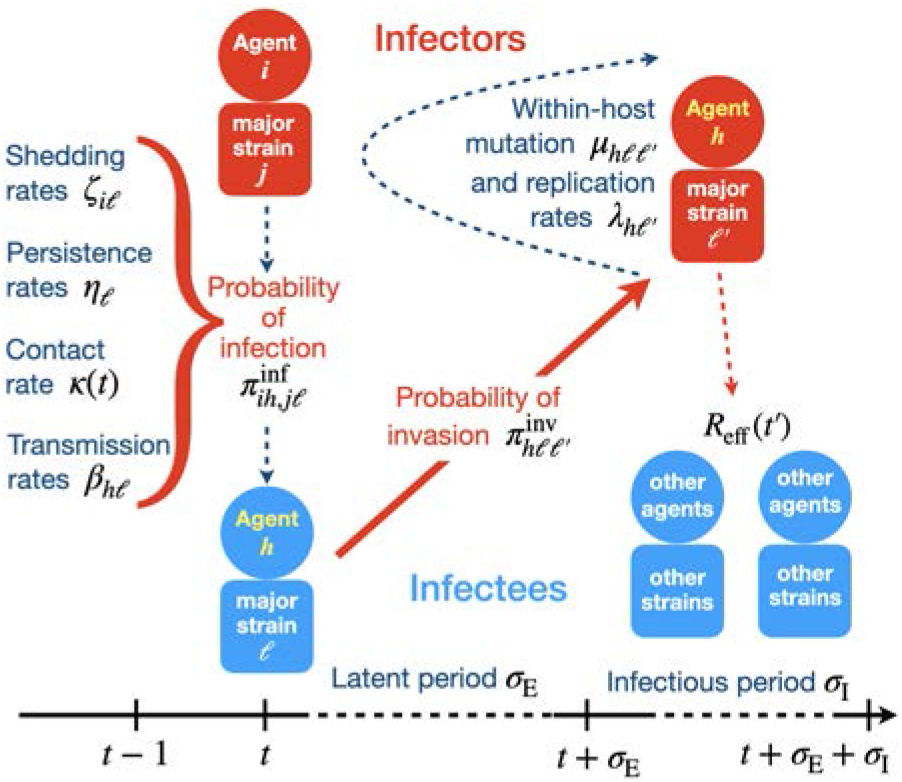
An overview of the processes included in our M-SEIR model (see Table 1 for equation references.) The probability 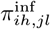 of A_*h*_ being infected primarily with pathogen *ℓ* in terms of receiving an effective dose from agent A*i* is computed in terms of a concatenation of shedding rates (*ζ* _*iℓ*_), environmental persistence rates (*η*_*ℓ*_), and host transmission (*β*_*hℓ*_) processes (SOF Eq. A.12) and includes both waning and cross immunity factors. The probability 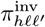 that the dominant variant emerging in host A_*h*_ is variant *ℓ′* given initial infection with variant *ℓ* is computed in terms of within-host mutation and within-host replication process (SOF Eq. A.13) and also includes both waning and cross immunity factors. These two probabilities are then used to compute the overall probability *π*_*ih,jℓ*′_ (SOF Eq. A.14) that infector *i*, infected with major variant *j*, infects infectee *h* with major variant *ℓ′*. The quantity *R*_eff_ (*t′*) is the expected number of individuals each infectious agent is expected to infect around time *t′* ∈ [*t* + *σ*_E_, *t* + *σ*_E_ + *σ*_I_], where *R*_0_ = *R*_eff_ (0) is estimated for our model using SOF Eq. A.26.

The reason for our inclusion of an adaptive contact rate process is that the local nature of contact rate patterns is well established as an important driver of outbreak dynamics [15]. If contact rates remain unchanged during the course of an epidemic, then a classic incidence curve (as in Fig. 2) will be the result. However, repeated peaks associated with consecutive outbreak waves arise as a results of implementing and then relaxing social distancing measures [15]. In the absence of social distancing drivers, which vary greatly from one location/region/country to another, an automated way to evaluate the effects of social distancing measures is through an adaptive contact process of the type that we include in our M-SEIR RAMP.

**Table 1:** Variables, indices and functions in our M-SEIR RAMP

| Symbols | Variables and indices | Equation<br>(See SOF) |
| --- | --- | --- |
| **Variables** |  |  |
| $t$ | time (variable and function values depend on time) | |
| $N_S, N_A, N_D$ | size of sets **S**, **A** and **D** | Eq. A.1 |
| $J, j, m, \ell$ and $\ell'$ | variant entropy and indices $(0, \dots, 2^J - 1)$ | Eq. A.2 |
| $N_I, N_{I_j}$ | total and variant $j$ infectious class size | |
| $A_i, A_h$ | specific agents $i, h = 1, \dots, N_A(t)$ in set **A** | Eq. A.4 |
| **Functions** (if a parameter now it may be elaborated later as a function) |  |  |
| $\kappa$ | adaptive contact rate | Eq. A.7 |
| $\omega_{ij}$ | waning immunity of $A_i$ w.r.t. variant $j$ | Eq. A.6 |
| $c_{mj}$ | cross immunity encountered by variant $j$ when agent previously infected with variant $m$ | Eq. A.8 |
| $\phi_{ij}$ | immunity modifier | Eq. A.8 |
| $\zeta_{ij}$ | shedding rate of variant $j$ by infector $A_i$ | Eq. A.9 |
| $\eta_\ell$ | environmental persistence | Eq. A.10 |
| $\beta_{h\ell}$ | variant transmission to infectee $A_h$ | Eq. A.11 |
| $\pi_{ih,j\ell}^{\text{inf}}$ | probability $A_i$ infects $A_h$ | Eq. A.12 |
| $\mu$ | mutation process factor | Eq. A.13 |
| $\lambda_{\ell'}$ | within-host replication rate | Eq. A.13 |
| $\pi_{h\ell\ell'}^{\text{inv}}$ | probability $\ell'$ is major variant when $\ell$ invades | Eq. A.13 |
| $\pi_{ih,j\ell'}$ | probability $\ell'$ is major variant in $A_h$ when $j$ is major variant in $A_i$ | Eq. A.14 |

**Figure 2:**
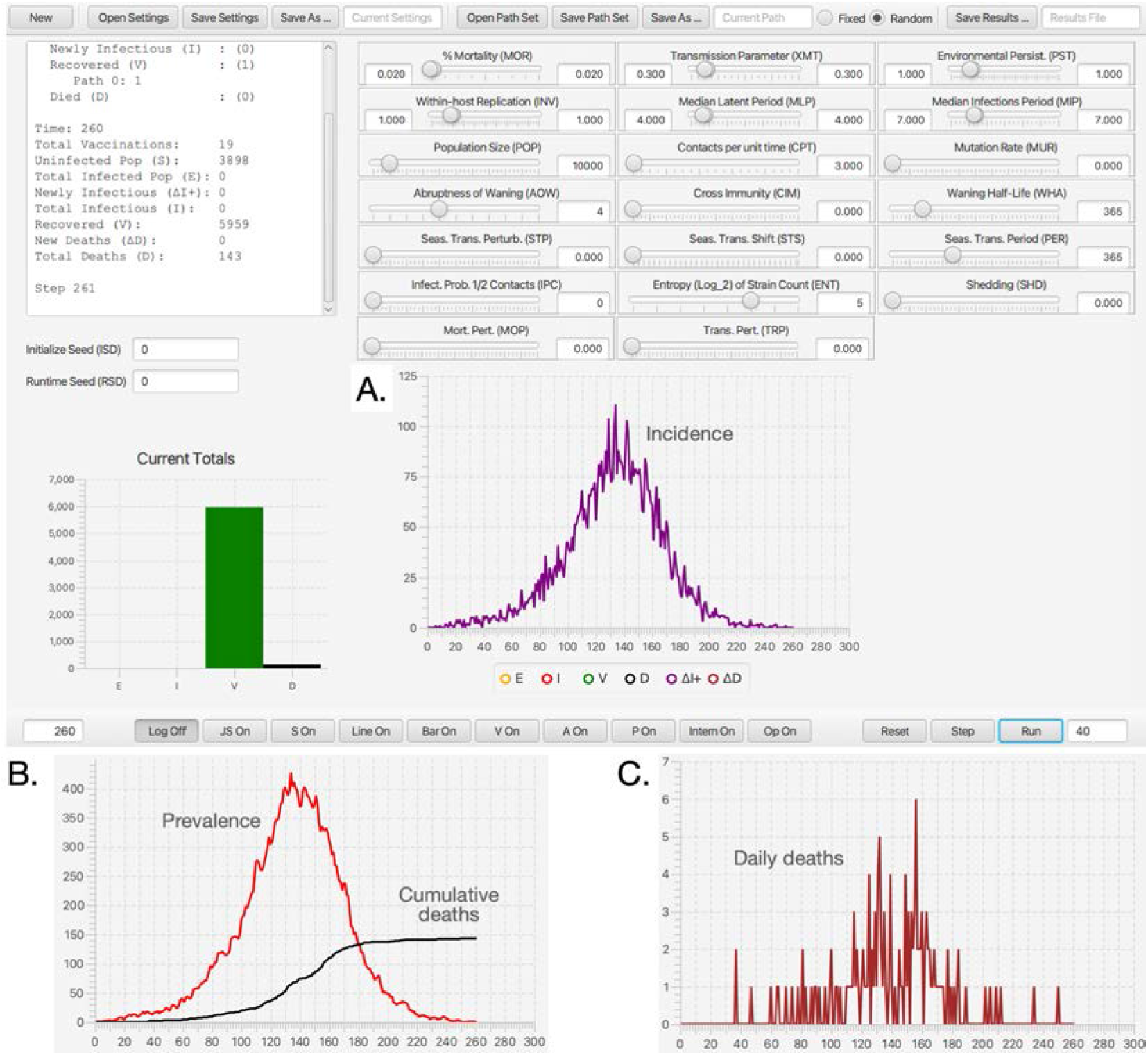
**A**. The dashboard of our Java Runtime-Alterable-Model Platform (RAMP) SEIVD (S=susceptibles, E=exposed, I=infectious, V=immune, D=dead) individual-based model (IBM) and simulations obtained using the parameters values depicted in the slider windows (also see Table 2). The top left window of this dashboard contains information on the final state of the population (in this case *S* = 3898 and *D* = 143 in a population of *N*_0_ = 10, 000), the bottom left bar graph of dashboard panel is the final values of E, I, V and D at epidemic cessation at time *t* = 166 (days) or the simulation run time, whichever comes first. Dashboard also shows a graph of incidence (purple: selected using colored buttons below the graph). The bottom ribbon of the dash board has a series of radio buttons that respectively open a Log, a JavaScript (JS), and a Scripting (S) window, Line and Bar graph windows (for multivariant runs), as well as windows for controlling vaccination strategies (V), listing realtime agent information (A), pathogen parameter values (P), monitoring probability computations (Intern), coding and controlling runtime alternative operations (Op), and three runtime buttons (Reset, Step, Run). **B**. Graphs of prevalence and cumulative deaths (cut out from main panel when only the red and black buttons are on) and **C**. daily deaths (crimson button) are pasted below the dashboard.

To illustrate the application of our M-SEIR RAMP, we used it to explore aspects of disease incidence and prevalence profiles using parameters that are applicable to the SARS-CoV-2 pathogen at the start of the COVID-19 pandemic. For example, we compare constant and adaptive (viz., prevalence dependent) contact rate processes under different waning immunity scenarios. We also explore the emergence of variants for different mutation and variant transmission rates. Additionally, we show how our M-SEIR RAMP can be used to evaluate the efficacy of uni- and multivalent vaccines applied at various time-dependent rates, where choice of valency may switch in response to realtime monitoring and surveillance data. Such adaptive vaccination programs may be required to combat the evolutionary arms race between vaccine efficacy and the evolution of new pathogen variants [25, 28, 29]. We hope, however, that our results and subsequent investigations using our M-SEIR RAMP provide the kinds of quantitative analyses that can help formulate highly effective local or country level vaccination programs that avoid some of the vaccination rollout pitfalls revealed by our analysis, as well as encourage the adoption of effective adaptive vaccination programs.

**Table 2:**
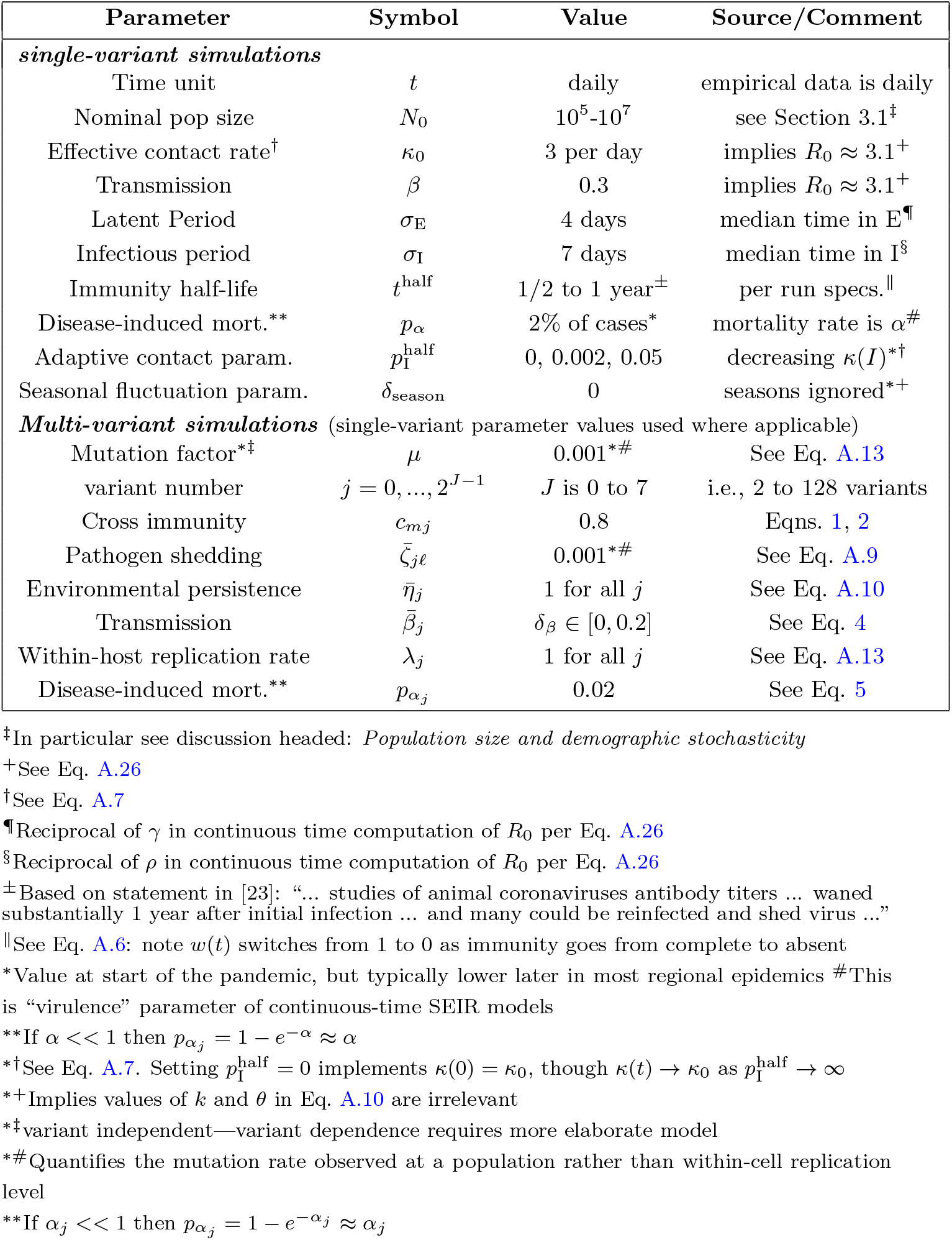
Parameter values used to simulate single and multivariant outbreaks

## 2 Materials and Methods

### 2.1 Our M-SEIR in a nutshell

We constructed an individual-based model (IBM) of a susceptible-exposed-infectious-recovered (i.e., an SEIVD model, where removed R are split into V=immune/vaccinated, and D=dead) epidemiological process [30, 31] in a homogeneous population with a random encounter contact rate parameter *κ*_0_ > 0. Our formulation allows for the emergence of multiple variants of the pathogen during a concatenation of process depicted in Figure 1 and listed in Table 1. Specifically, our formulation includes a host immunological waning process [23, 32] and a mutational process that impacts both transmission of mutant variants from the infectee and genetic drift [11, 24, 33] of variants within the infector, with rates impacted by cross immunity effects. We also allowed for variation in pathogen variant transmissibility (i.e., in the *β* > 0 parameter of the frequency dependent transmission function *βSI/N* [34, 35]) and pathogen virulence as represented by the disease-induced host mortality rate in the sense of Anderson and May [36] (and often represented by a parameter *α* ≥ 0 [34]).

The detailed formulation of our model and its algorithmic implementation is provided in Appendix A (SOF), with references to relevant equations in this provided in Table 1. In a nutshell we:

1. defined a set of 2^*J*^ pathogen variants (user selected value for variant entropy *J* ranging from 0 to 7; pathogen index *j* = 0, …, 2^*J*^ − 1) with a genetic-relatedness topology of a *J*-dimensional unit cube—i.e., each pathogen has *J*-loci that can take on one of two allelic values at each locus with immediate neighboring variants differing from each other by exactly one allelic value (0 or 1) at only one of the *J* loci
2. defined a population of *N*_0_ hosts as belonging at time *t* to either an epidemiologically naïve set of susceptible individuals **S** of size *N*_S_(*t*), a set **A** of *N*_A_(*t*) identified agents *A*_*i*_ (*i* = 1, …, *N*_A_(*t*)) whose epidemiological histories are known, or a set **D** of *N*_D_(*t*) individuals that have died from the disease
3. allowed pathogen variant-specific transmission “force” 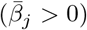 and virulence (*α*_*j*_ ≥ 0) parameters to vary in value among one another within a defined range 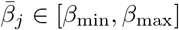 and *α*_*j*_ ∈ [*α*_min_, *α*_max_]
4. kept track of the total prevalence *N*_I_ as the sum of the prevalences of the individual variants 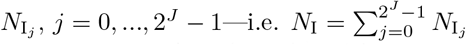
5. introduced a random contact rate function *κ*(*t*) with a constant parameter *κ*_0_ that is Poisson distributed on [*t, t* + 1), *t* = 0, 1, …, multiplied by an adaptive response function that reduces the contact rate with increasing disease prevalence, such that the *κ*(*t*) is reduced to *κ*_0_*/*2 when the 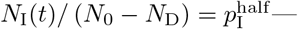 see SOF Eq. A.7
6. updated the epidemiological state of the agents A_*i*_ with respect to each of the variants *j* = 0, …, 2^*J*^ − 1, where the state with respect to particular variant *j* at time *t* is represented by a list that includes the following *J* entries pertain to the state of A_*i*_ with respect to pathogen variant *j* = 0, …, 2^*J*^ − 1. If the *j*^*th*^ entry is:
  a. 0, then agent A_*i*_ has never been infected with this variant
  b. E_*j*_(*t, τ*_*ij*_), was infected at time *τ*_*ij*_ ≤ *t* with this variant, but is not yet infectious for an expected period of σE units of time
  c. I_*j*_(*t, τ*_*ij*_), then agent A_*i*_ was infectious at time *t* with this variant, for an expected period of σI units of time, having been most recently infected (reinfections with the same variant may occur) with this variant at time *τ*_*ij*_ < *t*
  d. V_*j*_(*t, τ*_*ij*_), then agent A_*i*_ has now recovered from its most recent infection at time *τ*_*ij*_ with this variant and has some level of waning immunity to it
7. assumed that agent A_*i*_ can be infectious at time *t* with at most one dominant variant (denoted by the index *j*), although due to mutational processes this agent may infect other agents with variants related to this dominant variant at much lower rates (i.e., through application of a mutation factor *µ <<* 1, applied in our basic model through Eq. A.13)
8. assumed that agent A_*i*_ will have different levels of waning immunity to all of the variants to which it has been infected in the past
9. included waning immunity functions *ω*_*ij*_(*t*) (symbol is omega: SOF Eq. A.6) used to compute the level of immunity that agent A_*i*_ has to its most recent infection by variant *j*
10. included cross immunity effects (a *J* ^2^-matrix *C*) that apply both to the *infector* transmitting the pathogen and the *infectee* being invaded (inv; its “airport code” as described in SOF Fig. B.3) by the pathogen of interest, both of which reduce the likelihood of infection and variant drift by variant *j* compared with closely related variants *ℓ* (for a simple example of the matrix *C*, see Eq. 1 in Section 3 below)
11. computed an *infection probability* 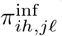 that agent A_*i*_ infected with variant *j* infects agent A_*h*_ with variant *ℓ* in terms of a concatenation of infector viral shedding (*ζ* _*iℓ*_; for a simple example see Eq. 6 in Section 3 below), viral persistence in the environment (*η*_*ℓ*_), and viral transmission (*β*_*hℓ*_) processes (Fig. 1)
12. computed an *invasion probability* 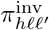 that an agent A_*h*_ infected with variant *ℓ* becomes infectious with variant *ℓ*′ as its major variant, in terms of the multiplicative effects of viral mutation (*µ*) and replication (*λ*_*ℓ*_) processes ongoing within an infectee A_*h*_ during this infectees exposed 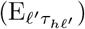 and infectious 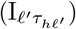 periods (Fig. 1)
13. computed the overall probability 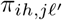 that an infector A_*i*_ infected with major variant *j* results in an infectee A_*h*_ expressingℓ′ as its major variant
14. assumed that waning and cross immunity to a particular variant is the same for both natural infections and vaccinations that use the antigen associated with that variant (of course we can easily extend our model to remove this assumption once data become available to support different waning and cross immunity rates for natural infections and particular vaccines)
15. implemented a discrete time individual-based stochastic SEIVD (here V represents individuals that have either recovered from infection or have been vaccinated, D represents cumulative dead; also see [37]) multivariant model that includes specifiable time-dependent univalent and multivalent vaccination implementations

### 2.2 Our RAMP implementation

Models of systems process can be coded as singular implementations model formulations using: i) highly efficiently compilable computer languages (e.g., C++, FORTRAN, Java); or ii) less efficiently, but more easily coded, scriptable (e.g., JavaScript, Python, Perl) computer languages. More conveniently and expeditiously, they can be coded up, as discussed in [38], using a systems modeling platform, such as Matlab’s SIMULINK, Mathematica, Stella, AnyLogic, Numerus, or Berkeley Madonna. Advantages of the latter include more rapid and accurate model development, though simulations may be slowed down by platform overhead. Between these extremes, we propose a more general approach to specific classes of systems’ models, where the basic system structure is fixed, but implementation of some elements can be easily and safely altered so that optional implementations are presented at runtime. We call such a design *runtime alterable-model platforms*. (RAMPs); and here we present a Java RAMP implementation of the M-SEIR described in the previous subsection.

The characteristics we envision for a model platform to be a RAMP are:

1. RAMPs include a set of model parameters (constants) whose values can be selected or specified (sometimes within a predefined range of values) at simulation runtime using a switch, nob, slider, or text-entry window accessed via a platform graphical interface or dashboard (e.g., see Fig. 2 and Table 1 which apply to our M-SEIR RAMP).
2. RAMPs include a specific set of *runtime alternative modules*, (RAMs), where the original can be redefined in a graphical interface window, and the unaltered original and the alternative routines are stored as a (preferably open-ended) numbered set. The original or any one of the alternatives can be selected for use at runtime (for a list of functions in our RAMP see Table 2).
3. RAMP implementations also provide an API for both remote and on-board scripting. This API enables control of all user aspects of the simulation, including the parameter set, run management, RAM options, and data retrieval. Script logic can alter parameter settings and RAM options as the simulation progresses. A Nashorn-based Javascript interpreter enhanced with API methods is provided.
4. The API can be accessed remotely using operating system facilities by external applications running concurrently with the simulator. Of particular interest is the ability to control the simulation from the R statistical platform. An R routine can be formulated to both manage the simulation run and to retrieve and process the resulting data. The RAMP simulation becomes a “virtual package” to the controlling R logic. See SOF Appendix B.

We implemented our RAMP using Java and made ample use of all of the features described above. Use of the RAM facility permitted experimentation with the several versions of cross immunity (e.g., Eqs.1 and 2). A Javascript program was used to control an adaptive vaccination strategy. A small R package serving as a driver was used in an R program that ran the simulations multiple times, extracted results into R data structures, and produced graphs showing statistical mean and standard deviation. More details on the graphical structure and implementation of our M-SEIR are provided in the presentation of both the results below and information in SOF Appendix B.

### 2.3 Simplifications and Running the model

In the examples presented in the next section, we have not taken advantage of the full complexity of the model. Thus, for example, in our multivariant simulations we have assumed that all variants are shed at the same relative rate (i.e., *ζ* _*ij*_ = 1 for relevant *i* and *j* = 0, …, 2^*J*^ − 1), have the same environmental persistence properties (i.e., *η*_*ℓ*_ = 1 for allℓ= 0, …, 2^*J*^ − 1), the same within host replication rates (i.e., *λ*_*ℓ′*_ = 1 for all *ℓ*′ = 0, …, 2^*J*^ − 1), and are all equally virulent (i.e., *α*_*j*_ = *α* for all *j* = 0, …, 2^*J*^ − 1). Obviously, these assumptions can be relaxed once suitable data are available for a particular pathogen to support variant specific shedding, persistence, within-host replication, and virulence values.

Further, in the absence of the kind of cross-immunity data obtained from cross-neutralization studies to be able to fit values to the cross-immunity parameters *c*_*mj*_ in the cross-immunity matrix **C** we contrast the following two cross-immunity scenarios with respect to a global cross-immunity constant *c* ∈ (0, 1). The first we call *cascading* cross-immunity since the level of cross-immunity diminishes multiplicatively with genetic distance of the two strains: viz.

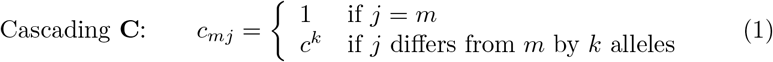

The second we call *escaping* cross-immunity since when the final allele in the array of J loci mutates from 0 to 1, it escapes completely from cross neutralization effects with all strains that have the original allele at the *J*^*th*^ locus: viz.

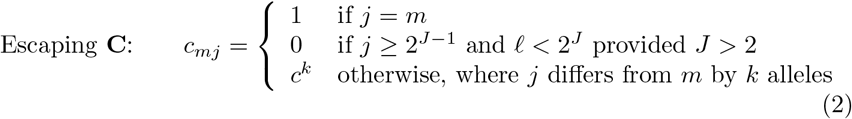

Obviously, this is an idealization of the *escape mutation* phenomenon, which we set up here to enable us to evaluate behavior of such mutations. For the purposes of this paper, idealized escape mutations are defined as those whose level of cross-immunity with the variants from which they arise is 0 (in reality some small level of cross immunity may remain).

Also, in the absence of comprehensive data that allows us to use realistic estimates for the relative transmissibility *β*_*j*_ and virulence *α*_*j*_ of various variants *j* = 0, …, *J*, we employ the following scenario facilitating formulations. These permit us to investigate the potential impacts of increased transmission and virulence with the emergence of new strains based on the number of mutations *d*_*j*_ needed to get from variant 0 to variant *j*. Specifically, for

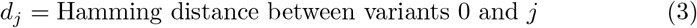

and for transmissibility and virulence perturbation parameters *δ*_*β*_ and *δ*_*α*_ respectively, we define

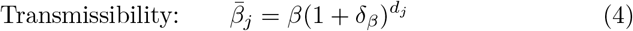

(the bar notation here reminds us that this value is used in the computation of *β*_*ij*_ according to equation Eq. A.8) and

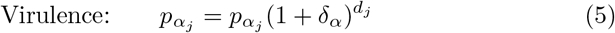

(this is a probability rather than a rate and we have to ensure *δ*_*α*_ is selected such that 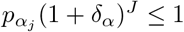.

Also for simplicity’s sake we assumed that infectee with major variant *j* will shed minor variants in the immediate neighborhood of *j* at comparative rate *ζ* ∈ [0, 1) and be major variant-independent: i.e., we assumed

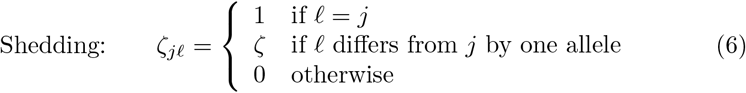

Finally, in this paper we will not investigate any seasonal effects, which is equivalent to setting *δ*_season_ = 0 in Eq. A.10, and using this setting in all our simulation.

The model itself can be accessed at Github, where instructions are available for launching and using our SEIVAgent application.

## 3 Illustrative Examples

### 3.1 Single variant simulations

#### Parameter values and baseline run

The first variable that needs to be determined is the unit of time we use for our simulations because all process rate parameters are scaled by its selection. Since the time resolution of empirical COVID-19 incidence and mortality data is daily, we selected our unit of time *t* to be days. Additionally, based on various sources including a metapopulation study of COVID-19 parameter estimates [39], we set the latent and infectious periods to be 4 and 7 days respectively. Basic SEIR epidemiological models do not separate out the processes of contact and transmission-per-contact, so we had some leeway on what values to choose for contact rates and transmission rates per contact because it is the value of the product of these that is important in determining the reproductive value, commonly referred to as “*R*_0_” for COVID-19. Hussein et al.’s [39] meta analysis of COVID-19 zeroed in on *R*_0_ = 3.14 as a mean value across a range of studies (95% confidence interval [2.69, 3.59]). By setting our baseline contact rate and transmission parameters to be *κ*_0_ = 3 and *β* = 0.3, we estimated the value of *R*_0_ in our model to be approximately *R*_0_ = 3.1. These and the remaining values of the parameters used in our simulations are summarized in Table 2.

#### Adaptive contact rate

None of the major outbreaks of COVID-19 around the world followed a classic “rise-and-fall” incidence curve because of social distancing and other measures taken to mitigate transmission once it had been determined that a full-blown outbreak was underway. These measures waxed and waned with government regulations and the perception that the outbreak was respectively under or out of control. This, in turn, resulted in incidence profiles that rose and fell multiple times (i.e., so-called waves) as these measures waxed and waned. Thus, it is not possible to replicate the incidence curves of any of the country/regional epidemics without characterizing the social distance and subsequent social relaxation measures driving their rise and fall [15].

In a general way, we can capture the gestalt effects of this kind of social behavior by assuming the contact rate *κ*(*t*) is influenced by current or recent prevalence levels, where current prevalence is given by the ratio of the number of infected individuals *N*_I_(*t*) to current population size *N* (*t*) − *N*_D_(*t*). Thus, in the various scenarios present below, we assume an adaptive response rate that has a maximum value *κ*_0_ when *I*(*t*) = 0 and is reduced to half this value, as a declining sigmoidal curve specified in Eq. A.7, when 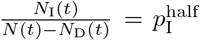. If we simulate the first year of an epidemic using our basic parameters (Table 2; also see parameter panel in Fig. 2) and adaptive contact half-max parameters for the cases Eq. A.7, when 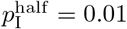 and 0.02 (i.e., 1% and 2% prevalence respectively), we obtain the percent of susceptibles (uninfected) and cumulative deaths by day 365 provided in Table 3.

**Table 3:** Basic runs with one million individuals (*N* = 1, 000, 000) using two different half-max adaptive contact parameter values 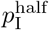 compared with listed countries^*^

| | $p_1^{\text{half}}$ | | USA | Italy | Czechia |
| --- | --- | --- | --- | --- | --- |
|  | 1% | 2% | (values under reported)<sup>†</sup> |  |  |
| **Uninfected at day 365** | 82% | 71% | 93% | 95% | 88% |
| **COVID-19 deaths by day 365** | 0.34% | 0.55% | 0.12% | 0.17% | 0.21% |

\*Data from Worldometer
<sup>†</sup>One year after the first 10 recorded cases in the countries concerned
<sup>‡</sup>Substantial under reporting occurs for both cases [40] and deaths [41].

For purposes of comparison, we also provide in Table 3 the percent of susceptible individuals and percent of deaths due to COVID-19 one year after the day on which more than 10 cases of COVID-19 were recorded to occur in the USA, Italy and Czechia (extracted from data provide at Worldometer). Since these data are known to be substantially under reported for both cumulative prevalence [40] and deaths [41], we felt that 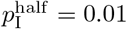 (i.e. 1% prevalence) provides a reasonable ballpark value for an adaptive contact rate half-max parameter for our various illustrations provided in below.

#### Population size and demographic stochasticity

Deterministic SIR/SEIR and related models will always produce an epidemic whenever the parameters ensure that *R*_0_ > 1 [2, 34]. Since these models do not include the demographic stochastic effects associated with finite populations, they are unable to capture the phenomenon of stochastic extinction of the epidemic before it gets going when a single infected individual is introduced into an otherwise infected population (with regard to the pathogen in question; see discussion in SOF A.4). In such models, results are either cited using percentages or numbers per thousand or per hundred thousand individuals and the actual population size is not regarded as a factor. Population size, however, is a factor in determining the absolute size of an epidemic once it gets started and deterministic models provided a robust assessment of the course of the epidemic in populations consisting of millions of individuals (other factors, such as spatial or contact network structure play a more important role than size per se [5–7]).

To get a sense of the effects of demographic stochasticity on populations of different sizes in our simulations, we compared the prevalence, incidence, and cumulative deaths obtained for single runs (runtime seed = 1) of a basic adaptive contact rate scenario (basic parameters plus 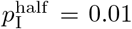) for cases where the initial population sizes where *N*_0_ = 10, 000, 100,000 and 1,000,000 (Fig. 3A-C). We also compared the mean prevalence plus/minus 1 standard deviation (sd) for 100 runs (runtime seeds ranging from 0 to 99) for the two cases *N*_0_ = 10, 000 and 100,000 over both the first year (Fig. 3D & E) and the first 100 day (Fig. 3F).

**Figure 3:**
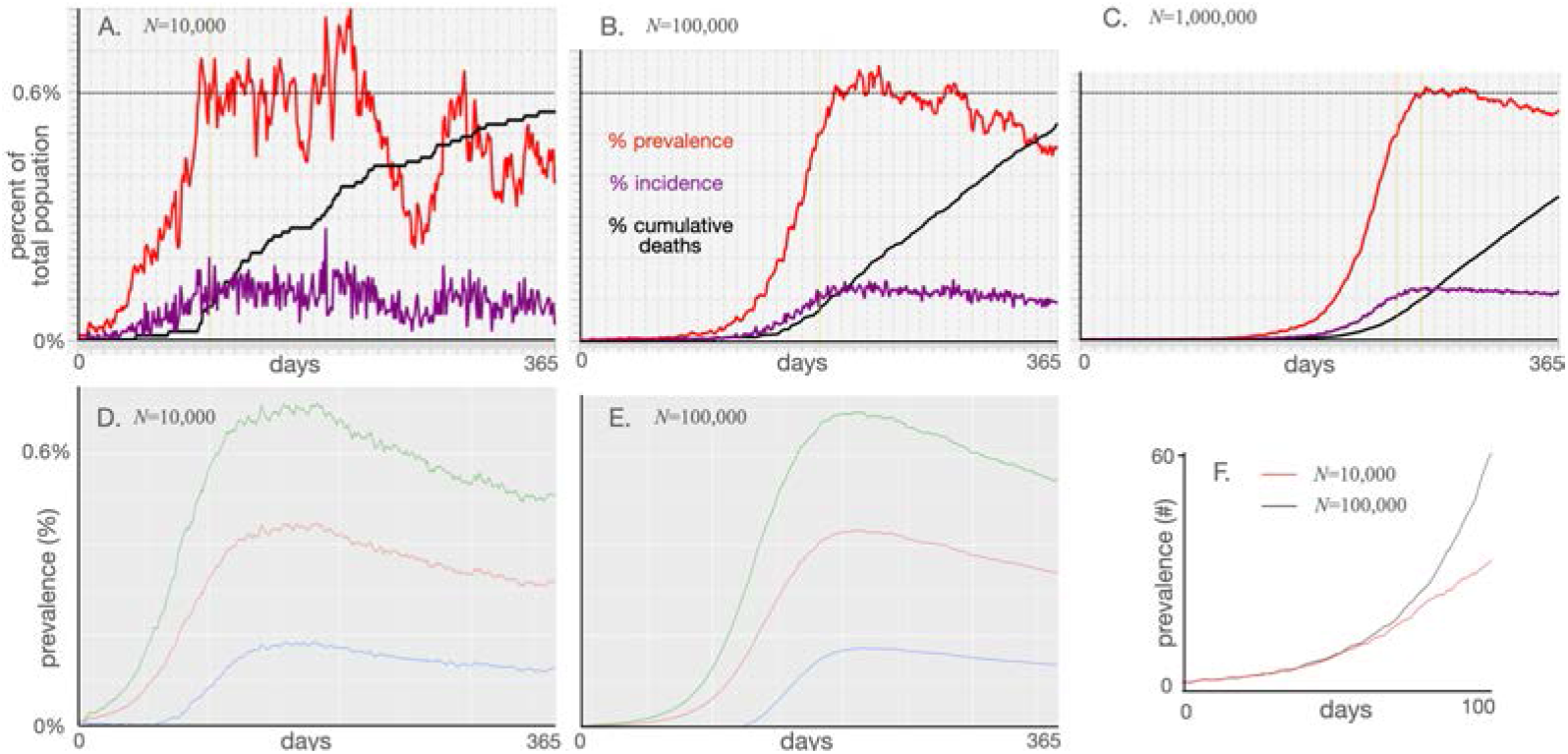
**A-C**: Plots of percentage prevalence (red), incidence (purple) and cumulative dead (black) for 365 day simulations using the parameter values given in Table 1 with the adaptive contact rate parameter 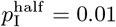 (see Eq. A.7) and *N* = 10, 000, *N* = 100, 000 and *N* = 1, 000, 000 respectively. **D-E**: Plots of mean percentage prevalence (red) over the first year plus (green) minus (blue) 1 standard deviation over 100 runs (runtime seeds going from 0 to 99) for the cases *N* = 10, 000 and *N* = 100, 000 respectively. **F**: Plots of the actual prevalence (number of individuals) for the first 100 days for the cases *N* = 10, 000 (red) and *N* = 100, 000 (black).

The results depicted in Fig. 3 can be encapsulated in the following four well-established principles:

1. The initial phase of an outbreak is independent of population size and establishment of an epidemic depends solely on the value of *R*_0_ (SOF, Append A.4). Thus, as we see in Fig. 3F, the first 50 days of the mean prevalence for the simulations of the cases *N* = 10, 000 and *N* = 100, 000 are virtually identical, only departing from one another from around day 50 onwards.
2. Beyond the initial phase, stochastic fluctuations are more evident in smaller than larger populations (compare Figs. 3A, B and C)
3. Ultimate prevalence levels, aside from stochastic fluctuations are independent of population size. Thus, for example, we see that prevalence maxes out at round 0.6% in all three cases (dotted line) across a range of two orders of magnitude in population size.
4. Mean population prevalence will always max out at lower levels than the prevalence reached in actual runs (viz. the max exceeds 7% individual runs in Fig. 3A-C while it is between 4% and 5% for the red curves in Fig. 3D&E) because the mean values take into account the fact a proportion 1*/R*_0_ of the runs go extinct within the first several weeks [42].

### 3.2 Multivariant simulations

We carried out a series of multivariant simulations with *J* = 4 (i.e., 16 variants can emerge) in a population of size *N* = 50, 000. We compared the scenarios of cascading cross immunity with *c* = 0.8 (Eq. 1) and transmissions rates the same for all variants (Fig. 4A) with the same cascading cross immunity as in Fig. 4A, but now we allowed transmission to increase by 20% for each mutation difference between variant *j* and variant 0 ((Eq. 1; *β*_*j*_ = 0.3, *j* = 0, …, 15, *δ*_*β*_ = 0.2 and *β*_*j*_, *j* = 1, …, 15, determined using Eq. 4). Finally, we compared the scenario of cascading cross immunity with that of escaping cross immunity for the case *c* = 0.8 (Eq. 2), and obtained the results provide in Fig. 4C. The severity of each scenario is encapsulated in the total death statistic over the course of the two-year simulation.

**Figure 4:**
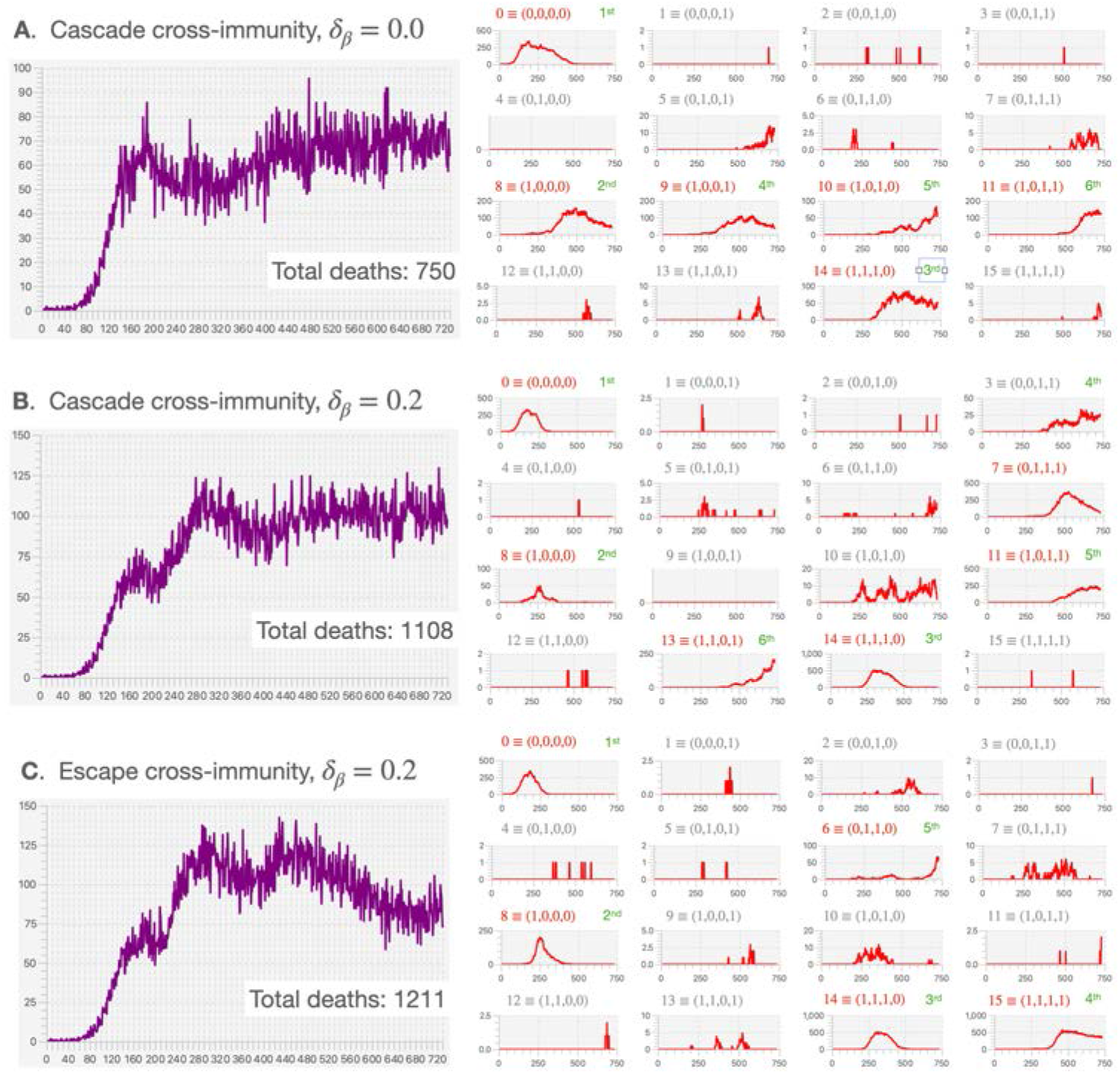
Total daily incidence (ΔI+: purple) and variant-specific prevalence (I: red) for a 16-variant epidemic in a population of size *N* = 50, 000 (for other parameter values see Table 2) are plotted over a two-year period for the three cases: **A**. cascade cross immunity *δ*_*β*_ = 0.0 (i.e., all *β*_*j*_ = 0.3, *j* = 0, …, 15) and *δ*_*β*_ = 0.2 (i.e., *β*_0_ = 0.3, *β*15 = 0.622 and *β*_*j*_, *j* = 1, …, 14, determined using Eq. 4). Variant number and corresponding binary representation as labeled in red for dominant or co-dominant variants (incidence at some point > 50 individuals per day) and gray for minor variants (incidence always *<* 50 over the two-year simulation). The order of emergence of dominant or co-dominant variants is labeled in green. Note that each panel has its own vertical scale but all plots are over 730 days (even in cases where the horizontal axis label go to 750).

Our primary observations comparing the results plotted in Fig. 4A-C and other runs (not shown here) of the same scenarios using different runtime seeds, are the following:

- In all three cases the initial variant, by construction, is 0 ≡ (0, 0, 0, 0). In our three scenarios, this variant was followed by chance by the emergence of variant 8 ≡ (1, 0, 0, 0), but this is common to all three scenarios because they use the same sequence of pseudo random numbers in their simulations. Using different runtime seeds, however, leads to other variants than 8 emerging to replace variant 0. Thus, the mutant identity (i.e., its binary representation) of the variant to first emerge is somewhat random, but it is going to be influenced by having different transmission values for each variant (scenarios B and C), as well as the possibility of an idealized escape mutation (scenario C).
- We expect variants that have an idealized escape mutation to emerge early, as is the case in scenario C in which variants 8-15 have the idealized escape mutation. In particular, in Fig. 4C, we see that the second to fourth variants to emerge all have the idealized escape mutation (i.e., variant 8 then 14 and then 15), and finally variant 6 ≡ (0, 1, 1, 0) emerges because of the cross-immunity between all variants with the idealized escape mutation finally comes into play.
- When *δ* > 0, the variants with the higher values of *β* come to dominate, though they take time to emerge. In our cascading cross-immunity case with *δ*_*β*_ = 0.2, the most transmissible of these (variant 15 ≡ (1, 1, 1, 1)) had yet to emerge within the simulated 2-year period. The existence of the idealized escape mutation, however, does facilitate the emergence of variant 15 at the beginning of the second year (Fig. 4C). Another run of this scenario (runtime seed=1; results not shown here) has variant 15 emerging very early (around day 120). Further, due to the effects of cross immunity, this variant was replaced by variant 11 ≡ (1, 0, 1, 1) around days 450-500. Variant 15, however, as result of waning and cross-immunity affects, reemerges again around day 600, with variant 11 declining over the last three months of the second year.

### 3.3 Vaccination rollout

#### Single valency vaccinations

As illustrations of potential issues associated with the design and implementation of vaccination programs, we first considered vaccinating individuals in a population of 100,000 subject to an epidemic involving a single variant of the pathogen. We note that in a population of *N* = 100, 000 individuals, a vaccination rate of *v*(*t*) = 0.001 involves vaccinating an average of 100 individuals per day with daily variation following a binomial distribution (i.e., a standard deviation of just under 10 individuals per day).

Rollout of our vaccination program began on day 366 after the start of the outbreak and ran for two years beyond that to day 1100 (Fig. 5). Such scenarios place us within the context of the COVID-19 epidemic since vaccinations were only available from around the second year onwards. In our first two scenarios, we selected individuals respectively at rates 0.1% (*v* = 0.001) and 0.2% (*v* = 0.002) of the population each day (Fig. 5A & B). Only individuals that had not been previously vaccinated were selected, but selection was otherwise random.

**Figure 5:**
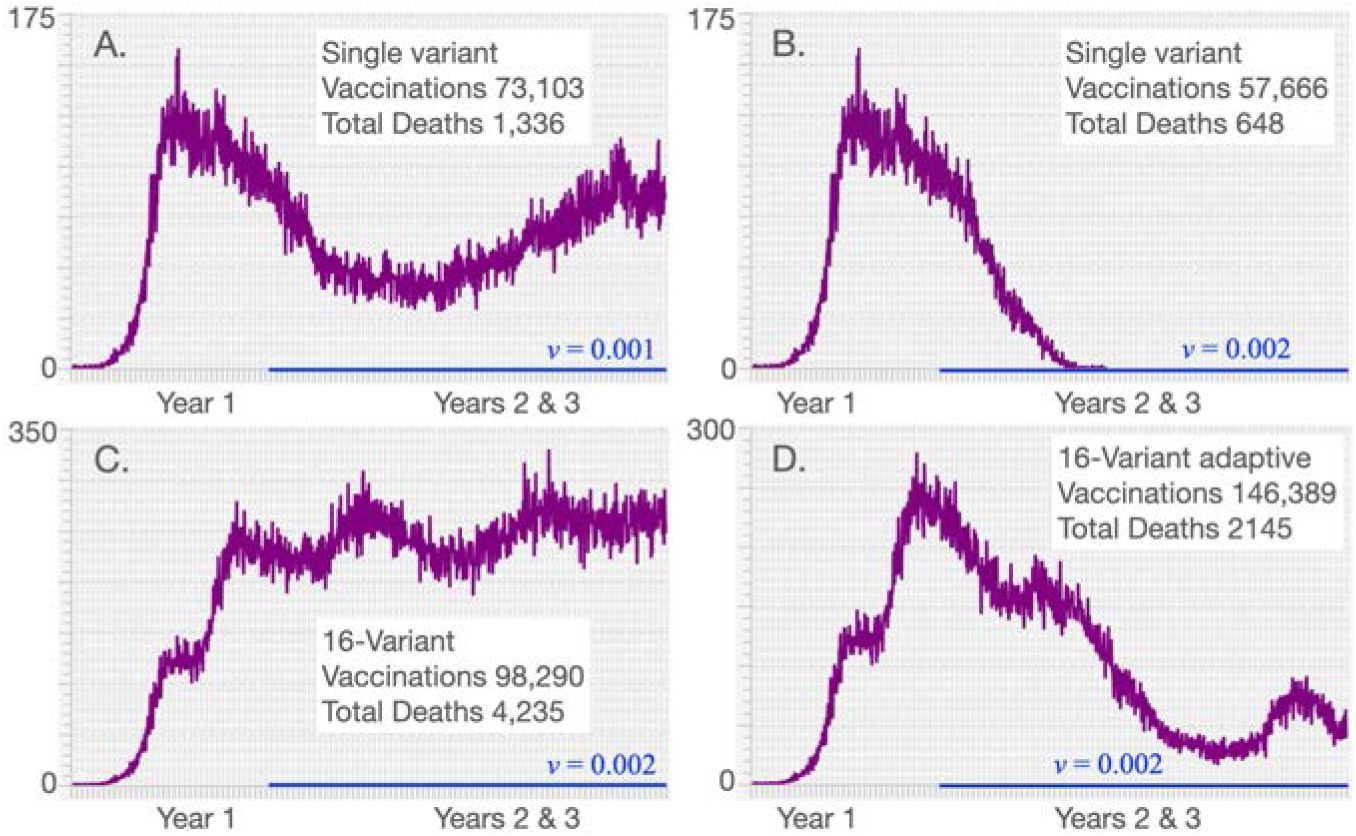
Incidence (ΔI^+^: purple) is plotted over 3 years for the baseline run (parameters given in Table 1 with *N* = 100, 000) for the cases where vaccination rates *v*(*t*) (indicated by blue lines) are applied during the second and third years only to individuals not previously vaccinated but otherwise selected at random (for clarification, the average number of individuals vaccinated each day is 100*v*(*t*)% with variation following a binomial distribution). Our first two simulations involve vaccination roll-out programs in a single variant epidemic at vaccination rates **A**. *v*(*t*) = 0.001 and **B**. *v*(*t*) = 0.002 (respectively 0.1% and 0.2%) of individuals not previously vaccinated, but otherwise chosen at random. Our second two simulations involve vaccination rollout programs in a 16-variant epidemic, both at vaccination rates *v*(*t*) = 0.002, involving **C**. individuals not previously vaccinated and **D**. a bivalent adaptive vaccination program in which previously vaccinated individuals could be vaccinated again with a new valency vaccine, as described in the text.

Additionally, we simulated a 16-variant scenario in which individuals were vaccinated at against variant 0 at rate *v* = 0.002 (Fig. 5C). Again, individuals were selected at random from the set of those that had not been previously vaccinated. By vaccinating individuals against variant 0, some immunity was conferred against variants 1-7 through cross-immunity relationships according to the Escaping **C** case with cross-immunity parameter *c* = 0.8 (Eqn. 2). In this scenario, variants 8-15 contain the idealized escape mutation.

Our focal question with regard to the first two scenarios was: What vaccination level is needed to extinguish the epidemic in the population encompassed by the vaccination rollouts for the populations concerned? From these two simulations (Fig. 5A & B) we see that vaccination rate *v*(*t*) = 0.001 was insufficient to eliminate the pathogen from the population, while *v*(*t*) = 0.002 was able to eliminate the pathogen within 10 months from the start of the vaccination rollout. Further, in the first of these simulations (Fig. 5A), we see a resurgence of incidence in year three, which implies that the effects of waning immunity in this case are essentially “outrunning the vaccination rate.”

Our focal question with regard to a comparison of scenarios two and three (Fig. 5B & C) was: Does the vaccination rate *v*(*t*) = 0.002, which was able to exterminating the outbreak in the 1-variant case, remain able to exterminate the outbreak in the 16-variant case when an idealized escape mutation is involved? The answer to this question from the observed incidence curve (Fig. 5C) is a resounding no. In fact, the total death rate over the three year period rose from 0.13% of the population (1336 individuals) to 0.42% of the population (4,235 individuals).

#### Adaptive bivalent vaccinations

With the emergence of new variants, the possibility exists to modify vaccines to contain or induce the production of antigens that directly target the variant in question (i.e., rather than through cross-immunity that arises when a related variant is the direct target) [28]. Further, it is possible for vaccines to be multivalent in terms of directly targeting more than one variant at time [28]. In our third vaccination scenario, a univalent vaccine applied at a rate *v* = 0.002 failed to bring a multivariant epidemic under control. Thus we were motivated to explore a scenario to see what would happen with a bivalent vaccine that was implemented adaptively in the sense of its two valencies following the two dominant variants.

In the specific vaccination rollout program that we employed in our fourth simulation, we did not account for logistical, production, and variant monitoring constraints. Such constraints, of course, exist and vary across locations and populations: in real applications, they need to be taken into account. The program we employed assumes that we are able to alter the valency of the vaccination used every 15 days, based on an ability to identify the two variants that are most prevalent at each of these successive 15-day-apart observation points (from day 365 to day 1085, which is the start of the last 15 day period ending just prior to the start of day 1100). If only one variant had an incidence exceeding 9 individuals on an observation day, then the vaccination applied over the next 15 day interval was monovalent for the dominant variant, otherwise it was bivalent for the two variants that had the highest incidence on that observation day.

As with the non-adaptive vaccination rollouts, individuals were selected at random from a pool that had previously not been vaccinated with the particular valency-specific vaccine (either bivalent or monovalent). However, in the bivalent vaccine case, if an individual had previously been vaccinated to only one of the two variants defining the latest vaccine, then these individuals were incorporated into the vaccination pool from which individuals were randomly selected for vaccination. If such individuals were selected then the start of the waning time relating to the previous vaccination was reset to start anew. Thus, with this program, it is possible for individuals to be vaccinated more than once.

The results of this simulation are depicted in Fig. 5D, where we see that this vaccination program is much more effective in preventing deaths than the monovalent variant 0 program applied to the same 16-variant epidemic at the same vaccination rate Fig. 5C (total deaths are 4,235 in the former versus 2145 in the latter case). The valencies of the vaccine applied during each new 15 day period are listed in Table 4. We note that the monovalent case involves considerably fewer vaccinations because of the “no revaccination with the same vaccine” restriction in our rollout program. In particular, over two years of vaccinating at a 0.2% rate per day, all individual are vaccinated in the case of Fig. 5C by day 859 while, in the case of Fig. 5, revaccinations kept occurring as individual that have not previously been vaccinated to one of the focal variants was revaccinated. Even in this adaptive rollout, however, the epidemic was only substantially lowered rather than completely extinguish. The latter event for the set of parameters used in our simulation requires a somewhat higher vaccination rate than 0.2% per day; or, perhaps it requires lower rates of waning immunity, higher cross-immunity rates, or the lack of an idealized escape mutation. All of these effects can be demonstrated through the selection of appropriate parameter values, but the specifics are only relevant when the model is applied in a real world situation.

**Table 4:** Valency of adaptive vaccination over the interval [365, 1100]

| Time | Valency |
| --- | --- |
| [365, 470) | (9,14) |
| [470, 530) | (13,14) |
| [530, 680) | (13,15) |
| [680, 740) | (15) |
| [740, 905) | (10,15) |
| [905, 1025) | (10,12) |
| [1025, 1070) | (12,15) |
| [1070, 1100) | (15) |

## 4 Discussion

The amount of structure and data needed in complex biological systems’ models, depends on the questions that these models have been formulated to address [13, 14]. In this paper, we steered away from making specific predictions— because universal solutions are not always locally applicable. Rather, we focused on gaining insights into incidence patterns that can be expected when contacts are adaptive rather than fixed, multiple variants may emerge (typically sequentially over time), and open versus adaptive uni- and multivalent vaccination programs are implemented to try to eliminate local pandemics. Analyses that incorporate more complexity in the hopes of attaining greater realism, such as adding heterogeneity related to age and spatial structures, as well as behavioral and social groups, require data that is specific to a local population (town, city, county, or small country, etc.) Such elaborations are only worth incorporating when the study relates to a real world system supported by adequate data (the latter related to the complexity of the question that needs to be addressed, as discussed elsewhere [13]).

Of course, additional structure can be added to address questions of general interest. One obvious issue relating to our study would be to obtain a better understanding of the role informational delays may play in producing the type of incidence waves that have observed over the course of the COVID-19 pandemic (and as we have modeled in [43]). Such delays would lead to contact rates containing a time-lag rather than depending only on current prevalence levels. We might also spend more time deconstructing the relative importance of such time delays versus the emergence of more transmissible variants in accounting for these waves.

Beyond gaining a deeper understanding of some of the mechanisms responsible for the incidence patterns observed among local epidemics of the COVID-19 pandemic, a second and primary purpose of our paper is to present our M-SEIR RAMP as a platform that others may use to address issues of concern to them in formulating policies to manage local COVID-19 epidemics. This also has the advantage of providing an exemplar of our novel RAMP (runtime alterable model platform) concept and the methods we used to construct it. At this time, the primary value of our M-SEIR RAMP itself may be in testing various vaccination strategies as they relate to variant emergence [44]. Clearly, such applications would require more specific variant-related information on the comparative transmissibility *β*_*j*_, virulence *α*_*j*_, shedding 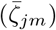, environmental persistence 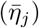, and within host replication rates (*λ*_*j*_) of newly emerging variants.

Equally important, though, in evaluating the impacts of vaccination strategies on local epidemics is obtaining variant-specific immunity and cross-immunity data. This includes waning rates, which we have not made variant specific. Our model, however, could be generalized to include variant specific waning rates represented by the parameter 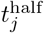 (Eq. A.6). It also includes information for characterization of the elements *c*_*jℓ*_ of the cross immunity matrix **C** (i.e., a generalization that renders Eq. 1redundant). Models are sorely needed to explore multivariant dynamics, particularly the epidemiological properties regarding shedding, environmental persistence, transmission, mutation, and within-host replication rates. These processes, acting together, determine the relative success of different variants and their actual impact on the severity of epidemics and the nature of vaccination programs needed to suppress them.

Making our model both location and variant specific could be undertaken using methods, such as Appropriate Complexity Modeling [13, 14], designed to enhance the relevancy of models. Further, in some cases it may be useful to add spatial or age-structure information to our M-SEIR or include a contact network [7], which itself may contain spatial or refined class category (e.g. age or work categories) information. In addition, our current implementation represents variant differences in terms of *J* loci with two alleles (denoted by 0 and 1 respectively) at each locus. A more realistic representation of the genetic basis of variant differences may involve genetic representations in which several alleles are possible at each locus. Further, the loci themselves may represent relevant molecular structures such as epitopes.

An advantage of our RAMP design features is that they provide a framework for elaborating or simplifying model details in the pursuit of different questions at various points in a pandemic. For example, suppose we are interested in pursuing inferences regarding the drivers of variant evolution at a various stages of the pandemic. We may first want to address questions relating to pandemic behaviour, driven by mutations that increase transmissibility. This is what actually happened with the appearance of the D614G and the alpha variants. A year or so into the pandemic, however, we may then want to explore processes that give rise to immunity-escaping variants. This, again, is what happened in reality. Our RAMP formulation gives us the flexibility to change the model part way through a pandemic. In particular, we can then test which among a set of alternative reinfection process is most likely to produce an escape mutation once reinfection begins to occur on a substantial scale. By configuring model drivers so that we first have a relatively simple evolutionary process and then switch to more complex evolutionary processes, our RAMP design facilitates comparing several competing explanations of observed patterns of variant emergence at different stages of a pandemic.

Although cross-immunity and immune waning are entangled in our immunity modifier functions (i.e., *ϕ*_*ij*_; see Eq. A.8), cross-neutralization data can be used to estimate the cross and waning immunity parameters using appropriate methods [45]. Such data are becoming more widely available through the application of improved serological and genetic methods [24, 46, 47]. Variant and cross neutralizing studies bring up a much neglected issue, which is the effect of dose (number of pathogens involved in the initial infection, also know as viral load) on the severity of the infection. Further, dose affects both the probability of host invasion (in the context of transmission), as well as mutational rates once host invasion has occurred. Effective dose differs from the questions of the number of vaccine doses (typically one versus two) versus the antigen or virus-like particle load in each dose [48]. In the context of vaccination, both these issues and the technology used to produce the vaccine [28] may well have an impact on waning immunity rates and cross-immunity values. Thus, the parameter values used in the model should ultimately be vaccine specific, once vaccine-specific waning data have been obtained.

In the coming years, as we obtain more information on the nature of waning and cross immunity to different variants of SARS-CoV-2, not to mention the vaccines as well, it will become more apparent to us whether or not COVID-19 will settle into global endemicity [32, 49] and require periodic vaccinations to combat new variants, as they arise over time. If this is the case, then constant vigilance and a well-designed vaccination program with respect to vaccinating the young and implementing booster vaccinations with appropriate variant valency will become the order of the day. Additionally, we anticipate extending our M-SEIR RAMP to include runtime alterable modules (RAMs) designed to compute the optimal time to administer vaccine booster shots of the same or different variant valencies. Implementation of these RAMs can play a decisive role in the rational design of effective and efficient COVID-19 vaccination programs worldwide. The need for efficacy is made apparent from the fact that our simulations suggest that it may be harder than currently anticipated to eliminate COVID-19 using non-adaptive vaccination programs.

Finally, our M-SEIR RAMP, with its RAMs, driver scripts and ability to be integrated with R and other software platforms and a JavaScript simulation driver window, provides the first example of a new concept in model implementation that facilitates model sharing and easy modification by users other than the original developers. We believe such platforms can come to play an important role not only in disease modeling, but in all fields of research that rely on models for comprehensive analyses of the behavior of systems of interest.

## Data Availability

Not applicable

## Acknowledgements and Funding Sources

This work was funded in part by NSF Grant 2032264 (PI: WMG).

## APPENDICES

### A Model Construction

Here we formulate an individual-based or agent-based (ABM) SEIR epidemiological model to include host immunological waning and pathogen genetic drift with variation across variant transmissibility and virulence and succintly refer to it as an elaborated SIR (M-SEIR) model. [50]

#### A.1 Assumptions, definitions, and states

The population consists of a well-mixed pool of *N*_0_ individuals that is homogeneous except for the fact that some are uninfected (denoted S), some currently infected (E: exposed and not yet infectious; I infectious and asymptomatic or symptomatic) or have been infected and are now either dead (D) or recovered/vaccinated with some level of immunity (V) to one or more of 2^*J*^ pathogen variants. This immunity wanes over time and its current level, augmented by specified levels of variant cross-immunity, factored into an agent specific time-dependent variant-resistance function that impacts the shedding of mutant variants by infectors and the within-host replication rates of mutant variants in infectees.

At the start of the epidemic, all individuals are assumed to encounter, on average, *κ*_0_ > 0 other individuals during each time period [*t, t* + 1], but this “effective contacts” rate adaptively decreases with increasing prevalence of the disease due to the implementation of non-pharmaceutical interventions (social distancing, hand washing, mask wearing, and other hygienic precautions). In our selection of epidemiological parameter values, a unit of time is taken to be a 24-hour day. Other scalings of time would then require appropriately adjusted epidemiological parameter values. Refined versions of the model could include age-related parameter values and contact rates, as well as contact tracing, quarantining, and isolation of infected individuals; but these will not be considered here.

Initially, at model time *t* = 0, all individuals are considered SARS-CoV-2 naïve susceptible apart from one individual who is considered to have just entered the infectious stage, infected by a pathogen designated as pathogen variant 0 (wildtype). Throughout the model simulation, the *N*_0_ agents in the population are partitioned into three disjoint sets: the set of SARS-CoV-2 naïve individuals, **S**(*t*), containing *N*_S_(*t*) (the susceptibles); the set of identified agents, **A**(*t*), containing *N*_A_(*t*) individuals who are either currently infected (time *t*) with a particular variant of SARS-CoV-2, or have some level of waning immunity to one or more variants of SARS-CoV-2; and the set of dead individuals **D**(*t*), currently of size *N*_D_(*t*). Only the individuals in **A**(*t*) are uniquely identified as they become infected for the first time and make the transition from set **S**(*t*) to set **A**(*t*), where they are sequentially labeled using the index *i* = 1, …, *N*_A_(*t*). The single infected individual at time zero will be designated Agent 1 (also known as patient zero and denoted by A_1_). Thus at time *t* it follows that

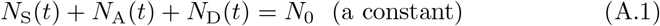

We note that individuals in set **A**(*t*) can be in a disease state E or I with respect to pathogen *j*, but simultaneously can be in multiple immune states if they have been infected with more than one pathogen variant in the past. We also note that the distinction between symptomatic and asymptomatic individuals in state I will not be considered here; and only need be incorporated if testing, quarantining, and treatment processes are included in the model.

The total number of pathogen variants is set by a parameter *J* > 0, where each pathogen is represented by a *J*-bit binary number. Thus, there are 2^*J*^ possible variants indexed by *j* = 0, 1, 2, …, 2^*J*^ − 1 where *j* is the decimal equivalent that corresponds to a given binary string. The initial variant, *j* = 0 is the binary string of *J* zeros.

Sets of stochastic epidemic events (i.e., transitions from classes S to E, E to I, I to V or D) are implemented at consecutive integer points in time (one set of events for each point in time). Events will only be considered on individuals that have been infected by at least one of the pathogens at some time after *t* = 0 (this means that initially the epidemic computation proceeds rather rapidly, but becomes more computationally intensive for each time step as time proceeds).

##### A.1.1 Pathogen set

At the start of the simulation (*t* = 0), the set of potential pathogens indexed by *j* = 0, …, 2^*J*^ − 1 is generated along with its associated environmental persistence 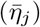, transmission 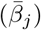, within host replication (*λ*_*j*_) and disease-induced mortality rate (probability of dying from the disease *p*_*α*_*j*) parameters. These may be specified or drawn from underlying distributions (e.g., the uniform distributions *β* ~`Uniform`[*β*_min_, *β*_max_] and so on). Also, our model includes two 2^*J*^ × 2^*J*^ matrices of constants that are associated with pathogen mutations during variant shedding (elements *ζ* _*jm*_) and cross-immunity (elements *c*_*mj*_) processes and thus involve but are conditioned on either the major variant that an infector is harboring or on immunological state of the agents involved. These are the shedding and cross-immunity matrices with elements *j, m* = 0, …, 2^*J*^ − 1, Thus we generate the following list of parameters associated with our 2^*J*^ pathogen variants:

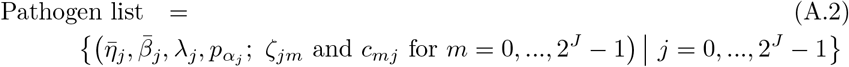

##### A.1.2 Agent states

In accordance with the above set of assumptions, each agent has the following basic disease states at time *t*, where disease states in agent A_*i*_ are referenced by the time *τ*_*j*_ > 0 at which the most recent infection with variant *j* has occurred (an individual may be re-infected after immunity to the variant has waned to relatively low levels):

1. S(*t*): An individual who at time *t* has not been infected with any variant of the pathogen up to time *t*. All these individuals belong to set **S**(*t*)
2. E_*j*_(*t, τ*_*ij*_): An agent A_*i*_ who was infected with variant *j* at time *τ*_*ij*_, but has not yet become infectious (this is an individual in the latent stage that lasts for *σ*_E_ units of time). All these individuals belong to set **A**(*t*)
3. I_*j*_(*t, τ*_*ij*_): An agent A_*i*_ who is currently infectious with variant *j*, after being infected with variant *j* at time *τ*_*ij*_ (this is the infectious stage that lasts for *σ*_I_ units of time). All these individuals belong to set **I**(*t*) ⊆ **A**(*t*)
4. V_*j*_(*t, τ*_*ij*_): An agent A_*i*_ who was infectious with variant *j*, having been infected with variant *j* at time *τ*_*ij*_, but is now non-infectious with regard to this variant—that is, recovered with some immunity to variant *j*, as well as some cross immunity to variants closely related to *j*. All these individuals belong to set **A**(*t*)
5. D(*t*): An individual at time *t* who has died after being exposed to and become infectious with some variant of the pathogen. In a refined version of the model, a record will be kept of the time of death and the variant that caused death. All these individuals belong to set **D**(*t*).

Since an agent A_*i*_ may be infected over time by more than one variant *j*, its complete epidemiological state is represented by a list

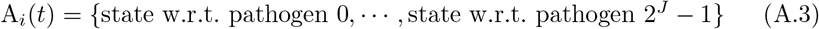

If a living agent does not fall into any of the categories 2 – 4 with respect to pathogen *j*, we denote its epidemiological state at position *j* as ∅ (the empty set). Consequently, if an agent A is susceptible at time *t* (i.e., an element of *S*(*t*)), then we write

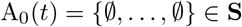

However, while such individuals are omitted from the A list (hence we did not subscript the agent A above), they may be recognized as “virtual members” with this implicit state. Some other examples are:

- If A_*i*_(*t*) is infected, but not yet infectious, with pathogen variant *j* at time *t* but has not been infected with any other pathogen in its past history, then

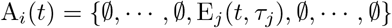
- On the other hand if A_*i*_ recovered from an infection with pathogen 0 at time *τ*_0_, and is now infectious with pathogen *j* at time *t*, having become infected with this pathogen at time *τ*_*j*_ then we write

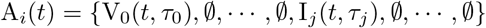

As we shall see, an agent history may contain at most one instance of either E_*j*_ or I_*j*_, while possibly containing multiple instances of V_*j*_.

##### A.1.3 Agent and index sets

At the start of each time period, we update the set of identified agents **A** by adding susceptibles that became infected with pathogens during the previous time period and removing agents that died during the previous time period. Thus if 𝕀_A_ is the index set for non-empty elements of **A**, with new indices added for newly infected susceptibles and indices removed for individuals that died, then by definition:

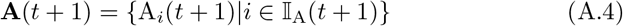

where the number of indices in the updated set 𝕀_A_(*t* + 1) is *N*_A_(*t* + 1) and the updated number of dead is *N*_D_(*t* + 1) at time *t* + 1.

For mathematical convenience all susceptibles S will also be referred to as A_0_:, i.e., there are *N*_S_(*t*) individuals referenced by A_0_ at time *t*. It will be useful to partition the set **A**(*t*) itself into three subsets at time *t* by identifying the sets **E**(*t*) and **I**(*t*) which respectively contain all agents that are currently in a state E_*j*_(*t*) or a state I_*j*_(*t*) at time *t* for some *j* = 0, …, 2^*J*^ − 1. We note the intersection of these two sets is empty—i.e., **E**(*t*) ∩ **I**(*t*) = ∅ —as will become apparent below from the transmission process rules set up below. We will use the notation

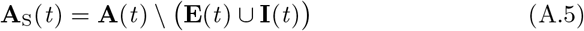

to denote the set of agents in **A**(*t*) but not in **E**(*t*) or **I**(*t*).

We also identify the set of infectious agents with infectious variant *j*. If A^*j*^ denotes an agent whose epidemiological state contains an entry I_*j*_(*t*, −), then

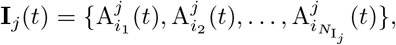

where the number of such agents is denoted by 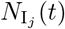, and its index set by

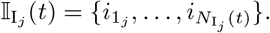

#### A.2 Epidemiological processes

##### A.2.1 Immunity

In compartmental SIRS and SEIRS models, a concept of waning immunity and its impact on epidemics is associated with the rates at which individuals in class R revert back to class S. In agent-based SIRS and SEIRS models, we have the opportunity to consider the immunological history of individuals and, hence, can take a more refined approach to the complex process of how pathogens in an infector A_*i*_ are passed on the an infectee A_*h*_. Here we model this as a probability generated from a concatenation of rates that include pathogen shedding by A_*i*_, the survival of pathogens in the environment, whether contained in feces, urine, sweat, mucosal secretions or water droplets excreted by an infector, and a process whereby pathogens gain access to a host (entering through wounds, mucosal membranes or other membranes in the pulmonary or alimentary systems). We then characterize pathogen within-host variant replication rates in terms of pathogen mutational and reproductive processes. The final outcome in our model is either host recovery with some immunity or host death. We also consider the induction of host immunity through vaccination and make the assumption that waning immunity is the same, whether it stems from natural infection or vaccination. Of course, these may be modelled in different ways should data become available to make this distinction an important modeling consideration.

###### Waning immunity

Recall that we use A_0_ to denote an anonymous (generic) member of **S** and that A_*i*_ for *i* > 0 refers to a specific individual with an associated state list/vector. If some specific A_*i*_ is in immune state V_*j*_ having been infected with this variant at time *τ*_*ij*_, we assume that the level of relative susceptibility of agent A_*i*_ to reinfection by variant *j* is given by (noting that the existence of the value *τ*_*ij*_ implies that infection of individual *i* by variant *j* at time *τ*_*ij*_ ensures that the V_*j*_ is no longer “null”)

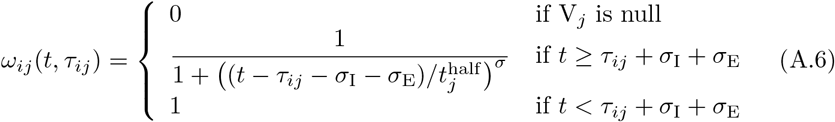

We note the following: 1.) the first case implies that *τ*_*ij*_ has yet to be defined; 2.) the second case is equivalent to the statement that *τ*_*ij*_ ≥ 0 now exists for variant *j*, since this occurs at time *t* = *τ*_*ij*_ (through the invocation of state E_*j*_(*t, τ*_*ij*_)); 3.) *ω*_*ij*_(*t, τ*) ranges from 1 (i.e. full “on”) at *t* = *τ* + *σ*_I_ + *σ*_E_ and decays to 0 as *t* > *τ* + *σ*_I_ + *σ*_E_ → ∞; 4.) agent *i* cannot be reinfected with its current major variant or with any other variant while it is currently itself in any state E_*j*_ or I_*j*_ for any *j* = 0, …, 2^*J*^ − 1; 5.) the larger the value of *σ* the steeper or more abrupt the switch is from full immunity (equal to 1) at time *τ* through 1/2 at time 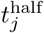 to approach 0 as *t* → ∞ (we set *σ* = 4 as providing an intermediate level of abruptness).

###### Vaccination

A vaccine may be designed to give immunity to one or more particular identified variant *j*. Vaccination strategies include vaccinating at a fixed rate *v*(*t*) (percent of individuals vaccinated at each time period) over a fixed period that begins at 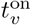 and ends at 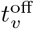 and can focus on drawing only on: i) individuals in the set **S**, ii) any non-infectious individual in **S** or **A**, or iii) any non-infectious, not previously vaccinated individual in **S** or **A**. The vaccine itself can be designed as follows:

- *Dominant variant vaccination at time τ*_vac_. An individual S or A_*i*_ vaccinated with the dominant variant, say *j*, at time 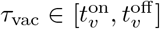 serves to add the disease state V_*j*_(*t, τ*_vac_) to that individual’s list. If the individual is already in state V_*j*_(*t, τ* ′) at time *t* > *τ*_vac_, then its status is updated so that at time *t* > *τ*_vac_ it is now V_*j*_(*t, τ*_vac_) rather than V_*j*_(*t, τ* ′)
- *Multivariant vaccination at time t*_vac_. An individual S or A vaccinated with a multi variant concoction at time 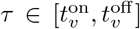, say with variant *j*_1_, …, *j*_*v*_, will have their disease status updated with regard to all these variants, as in the dominant variant case.

##### A.2.2 Infectious contacts

Infectious individuals are assumed to make 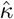 *effective contacts* each time period; where effective contacts are those that are sufficiently close and of a sufficiently long duration to constitute a “risk of transmission.” This rate is either a constant *κ*_0_, or in stochastic implementations drawn from a Poisson distribution with mean *κ*_0_, or in adaptive formulations (e.g., under social distancing behaviour) is a function of the severity of the ongoing outbreak. We also assume at time *t* that under a random contact process, proportion 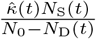 and 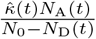 of these contacts will respectively be with susceptible and with uniquely identified agents, although only 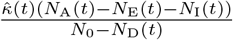 of those will be susceptible to infection with a new variant or reinfection with the same variant.

In the adaptive case, we assume *κ*(*t*) decreases from *κ*_0_ as the proportion of infectious individuals, *N*_I_(*t*)*/*(*N*_0_ − *N*_D_(*t*)), increases such that *κ*(*t*) = *κ*_0_*/*2 when 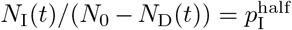. For convenience of implementation, however, we define the following “switching” (as apposed to hyperbolic) function

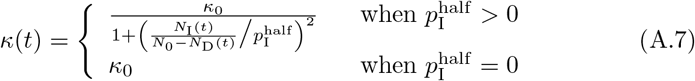

even though, from a continuity point of view, the top part of this expression implies that *κ*(*t*) → 0 at 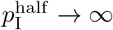.

##### A.2.3 Probability of infection

In deriving a probability 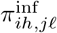 of an agent A_*h*_ being infected with variant *ℓ* by an agent A_*i*_ who is infectious with major variant *j*, we concatenate (i.e., multiply together) several process, each of which involves nominal constants. Thus, in all but one of these processes, the scaling of these constants can be normalized and given a relative set of values across variants though one set of constants though relative, will ultimately all be scaled by fitting the model to real data. In our treatment below, constants associated with shedding and persistence will be scaled while those associated with within-host replication will be kept unscaled to be ultimately fitted to data. In particular, the parameters 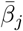 associated with pathogen transmission (i.e., from contact to the start of within host replication—see Fig. 1) will be scaled by fitting to epidemiological data, while the relative values for the different variants regarding pathogen shedding and environmental persistence can be fitted to experimental data collected to set values of these processes when considered on their own.

###### Pathogen shedding

We assume that shedding is affected by the immune state of the infector A_*i*_ and thus posit the shedding rates below for this individual when its major infectious variant is *j*. In general, we have a matrix of shedding rates 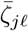 before accounting for immunity and cross immunity that is specific to agent A_*i*_. Immunity and cross-immunity act to reduce shedding rates through functions *ϕ*_*ij*_(*t*) ∈ [0, 1] that are computed in terms of A_*i*_’s waning functions *ω*_*im*_ with respect to variant *m* and a matrix of cross-immunity values *c*_*jm*_ that have been normalized so that *c*_*jj*_ = 1 for *j* = 0, …, 2^*J*^ − 1 and *c*_*jm*_ ∈ [0, 1] for *j, m* = 0, …, 2^*J*^ − 1. Specifically, we define agent-specific *immunity modifying functions*

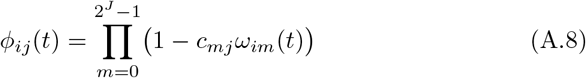

and assume that the shedding rates can be expressed as

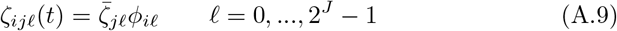

###### Environmental persistence

The persistence of pathogens in the environment are known to be impacted by humidity, temperature, airflow, and the surface properties of fomites [51]. This, and other factors relating the effects of weather on contact rates and efficacy, may result in overall pathogen transmission having a seasonal component to it [52]. In particular, viral persistence indoors may be much greater than outdoors, with a greater proportion of indoor contacts taking place during cold or wet weather. Thus the most appropriate place to introduce seasonal effects into epidemic processes is through contact rates and environmental persistence cycling over time with a period of one year (or even half-a-year if two comparatively spaced rainy seasons occur, as in some in tropical locations [53]) and an amplitude obtained by fitting parameters to data. Thus, in our model, we introduce constants 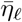, δ_season_ ∈ (0, 1)*k* (appropriately scaled, depending on the units of time) and *θ* and assume that

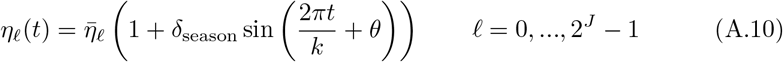

The case *δ*_season_ = 0 corresponds to constant values 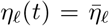 for all *t*, while if *δ*_season_ = 1 we get the largest possible fluctuation between 0 and 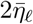. The constant *k* relates to the time units so we get one cycle per year, and *θ* shifts the cycle to set the points in time at which the maximum and minimum values of *η*_*ℓ*_(*t*) occur.

###### Variant transmission

In the context of a standardized dose (which will be modified by multiplying the variant *effective contact* and transmission by both pathogen shedding and environmental persistence functions), the differential rates of variant transmission, which we denote by *β*_*hℓ*_, will depend on a constant variant transmission rate parameter 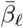 modified by a function that represents the immune state of the infectee at time *t*: viz., recalling SOF Eq. A.8

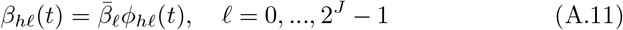

###### Probability of infection

Using a competing rates formulation [37] to compute the probability of infection as a concatenation of the process of infector shedding (*ζ*), environmental persistence (*η*) and transmission rates (*β*), we obtain

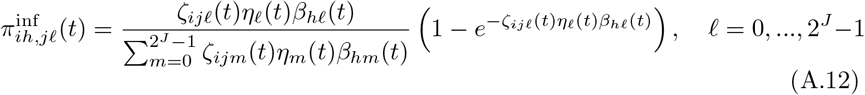

##### A.2.4 Within-host processes

If after receiving an initial infectious dose of pathogen, an individual is infected primarily with variant *ℓ*, then we expect this variant to dominate unless intrinsic mutational processes are high (which is not the case for COVID-19) or the individual has some immunity to this dominant variant. In the latter case the situation is ripe for an “idealized escape mutation,” that is one that evades the immune system completely, to arise.

If we nominally set the relative rate at which an individual invaded by variant *ℓ* has an infection dominated by variant *ℓ* (i.e.,*ℓ* in the terminology of [54] is the major variant of the infection) to be (1 − *µ*), then the probability that one of the other variants is *ℓ*′ ≠ *ℓ* is *µ* (in the case of COVID we assume that *µ* > 0 is very close to 0—e.g. of order 10^−3^ to 10^−6^—while for viruses lacking error correcting machinery it can be considerably larger and of the order 10^−1^). We can partition the latter probability according to a set of comparative variant within-host replication rates *λ*_*ℓ′*_, each moderated by its immune state function *ϕ*_*hℓ*′_ and a normalizing factor 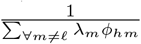 to obtain

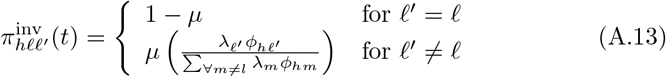

We stress that the parameter *µ* pertains to generating the probability for the transmission of mutants and is not an actual mutation rate for the virus (e.g., the host may have some mechanisms for removing most of the mutants before transmission of remaining variants occurs).

##### A.2.5 Pathogen progression equations

Probability that infector A_*i*_ with major variant *j* will result in infectee A_*h*_ express ℓ′ as its major variant is

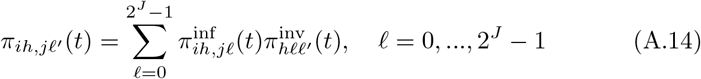

##### A.2.6 Single-variant case

In the single-variant case (*J* = 0), the waning immunity equation SOF Eq. A.6 reduces to (dropping the redundant index *j* = 0, and noting that the existence of a value *τ*_*i*_ implies A_*i*_ has been infected at time *τ*_*i*_ in the past)

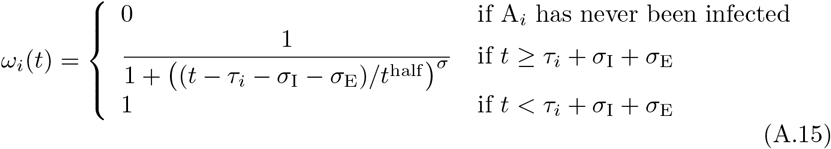

(recall we set *σ* = 4) and the modifying immunity functions *ϕ*_*ij*_ (SOF Eq. A.8) collapse to 1, which implies that the pathogen shedding functions 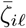 (SOF Eq. A.9) collapse to 1. Without loss of generality, we can also assume a single-variant value of *η* = 1 in SOF Eq. A.10, which implies that the probability of infection (SOF Eq. A.12) reduces to

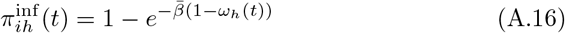

Further, since in the single-variant case there are no mutations to consider, it follows from SOF Eq. A.13 that 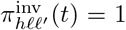 for all *h* and we finally have that 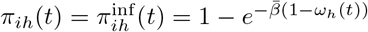 (SOF Eq. A.14) for all *h*.

#### A.3 Simulation algorithm

1. **Parameters selected at the start of a simulation**
  a. *N*_0_: Number of individuals in the population. Assumed to be fixed over time (i.e., the population is closed), but partitioned into sets **S, A** and **D** with respectively *N*_S_(*t*), *N*_A_(*t*) and *N*_D_(*t*) individuals in each set and satisfying SOF Eq. A.1.
  b. *J* : The log_2_ of the number of possible variants indexed by *j* = 0, …, 2^*J*^ − 1
  c. 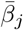 : variant dependent transmission parameters (the process between contact and the start of variant replication and nominally equivalent to transmission in SEIR models—see Fig. 1) for pathogen variant *j*
  d. 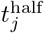 : The time it takes for immunity to variant *j* to have waned by half.
  e. 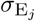 : The time it takes from initial infection for an infected individual to become more likely to become infectious than remain infected without being infectious.
  f. 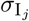: The additional time it takes beyond 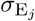 for an infectious individual to more likely transition beyond being infectious than remain infectious.
  g. 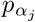 : The proportion of individuals leaving the infectious category that die, which implies that 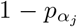 is the proportion that become immune.
2. **Initialization**
  a. Set up pathogen list (see Eq. A.2)
  b. Initialize the simulation by setting *t* = 0 and creating the agent list **A**(0) one infectious and *N*_0_ − 1 susceptible agents.
3. **Time** *t***: vaccination loop**.
  a. Carry out the vaccination process before going into the rest of the loops with the updated S and A sets after the vaccinations.
4. **Time** *t***: contact loop**. Set up contacts for the current round of encounters at time *t* (i.e., the inner agent-driven contact loop within the outer time-driven loop) and tag for outer loop update of disease status, as follows:
  a. *Numbers in various sets and associated index sets*. Identify the number of individuals *N*_S_(*t*), *N*_A_(*t*) and *N*_D_(*t*) in sets **S, A** and **D** at time *t* respectively, as well as the number of exposed (but not yet infectious) agents *N*_E_(*t*), infectious agents *N*_I_(*t*) and identified non-infected agents 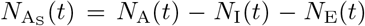. Break down the infectious agents tally into the number of agents 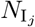 infectious with variant *j* = 0, 1, …, 2^*J*^ − 1. We will also need the index sets 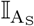 and 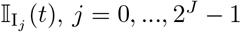 at time *t*.
  b. *Infectious contacts with each group*. The rate at which any individual contacts other individuals per unit time is given by the contact rate parameter *κ* > 0. Assuming random contact events over one unit of time, the actual number of individuals that agent A_*i*_ contacts at time *t* is then given by

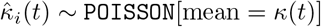

Of these, proportions

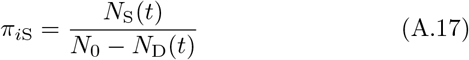

and

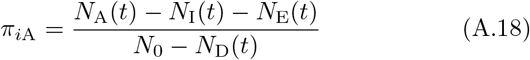

are expected to come from susceptibles in the sets **S**(*t*) and **A**_S_(*t*) (see Eq. A.5) respectively. Thus the actual number of contacts in set **S**(*t*), **A**_S_(*t*), and **E**(*t*) ∪ **I**(*t*) are

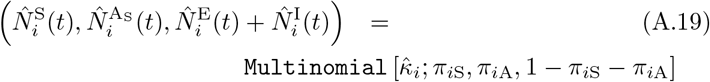

We note that only 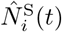 and 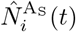 are of interest because individual in states E and I cannot be reinfected. Also, we make the assumption below that the first infection that an individual in set **A** contracts in this contact loop, is the one that counts (i.e., there will be no simultaneously infections with multiple variants). Finally, since contacting individuals is tantamount to sampling with replacement, the number of unique contacts (i.e., all multiple contacts are counted as a single contact) that agent A_*i*_ has with individuals in the set **S** is 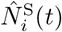 reduced by excluding multiple contacts (which under a random contact model is a negative exponential correction) to obtain

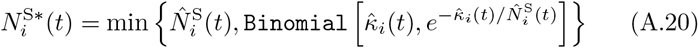

Thus if 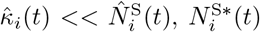 is expected to be very close to the upper value 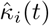. On the other hand, if 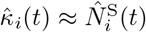, then 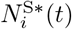 is expected to be around 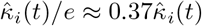. Additionally, after dealing with each agent *i* reduce in the size of *N*_S_(*t*) to take account of those agents that had been infected by agent A_*i*_ and had now entered the ranks of the set **A**.
  c. *Identify all infectious agents and their pathogens variants*. Among all agents in the set **A**(*t*) (Eq. A.4), identify those that have an infectious variant I_*j*_ for some *j* = 0, …, 2^*J*^ − 1. Thus, if the number of infectious agents with infectious variant *j* is 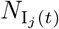 then consider the set

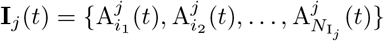

with index set

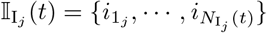

Initially, most of these sets will be empty, but will fill in over time.
  d. *Susceptible contacts*. The probability that an agent A_*i*_ with a variant *j* major infection infects a susceptible (nominally denoted by individuals of type A_0_) who then becomes infectious with dominant variant ℓ′ is given by the probability 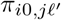 computed in Eq. A.14, which itself relies on expressions Eq. A.7–A.13. The actual number of individuals in the set **S** will make effective contact with one more infectious individuals is 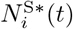 obtained using Eq. A.20. Thus, from a multinomial drawing, we can now generate the number of newly exposed individuals, 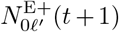 (the “+” is used to denote these are newly added and the “0” that they are coming from the set **S**), with major variant ℓ′ at time *t* + 1, have been infected by agent A_*i*_ with major pathogen variant *j* on the time interval [*t, t* + 1):

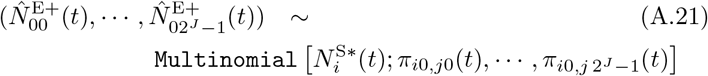

These individuals will be used to update list of currently infected individuals in the sets 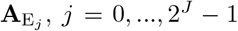 at time t+1, which is computed in the outer loop computation, as presented below. We also note that the probabilities in the above multinomial add to less than 1, so that at the end of the drawing a proportion of the individuals 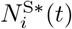 remain uninfected.
  e. *Agent contacts*. The number of agents 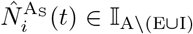 that come into contact with agent A_*i*_ over the interval (*t, t* + 1) is given by Eq. A.19. This number is drawn from the set 𝕀_A\(E∪I)_ with replacement and the following multinomial computation is used to determine how to update agent A_*h*_ at time *t* + 1 when coming into contact with agent A_*i*_ on the interval (*t, t* + 1) using the probabilities of transmission given in Eq. A.14. Specifically, agent A_*h*_ will become infected with major variant ℓ′ at time *t* + 1 is determined by the multinomial drawing

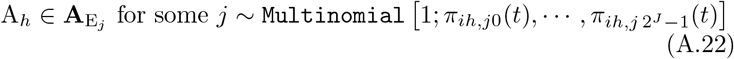

We note here that since the agents *A*_*h*_, *h* ∈ 𝕀_A\(E∪I)_ are drawn with replacement as the computation proceeds and the agents A_*i*_, *i* ∈ 𝕀 are cycled through, if a previously drawn *A*_*h*_ is drawn again, but has already been infected in the current round then we ignore the latest event, but keep the previous infection event intact. To obviate bias in this procedure, we need cycle through the agents A_*i*_, *i* ∈ 𝕀 at random rather than in numerical order.
5. **Time** *t***: disease progression loop**.
  a. *Individuals in* **A**_E_ *at time t*. An individual A_*i*_ ∈ **A**_E_ at time *t* and in state E_*j*_(*t, τ*_*i*_), *j* = 0, …, 2^*J*^ − 1, becomes either an individual in state E_*j*_(*t* + 1, *τ*_*i*_) with probability

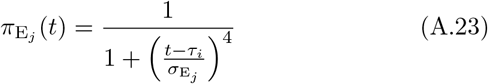

or transfers to state I_*j*_(*t* +1, *τ*_*i*_) with probability 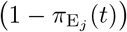 thereby entering class **A**_I_ at time *t* + 1.
  b. *Individuals in* **A**_I_ *at time t*. An individual A_*i*_ ∈ **A**_I_ at time *t* and in state I_*j*_(*t, τ*_*i*_), *j* = 0, …, 2^*J*^ − 1, becomes either an individual in state I_*j*_(*t* + 1, *τ*_*i*_) with probability

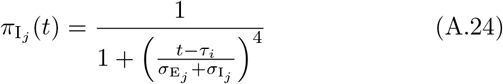

or leaves the set I_*j*_(*t* + 1, *τ*_*i*_) with probability 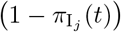. In this latter case, the individual either dies with probability 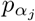 or enters the state V_*j*_(*t* + 1, *τ*_*i*_) at time *t* + 1 with probability 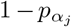 The total number of individuals dying over the interval [*t, t* + 1) is noted as having a value Δ*N*_D_(*t*).
6. **Time** *t* + 1**: outer loop update**. The outer loop records all the events that took place in the contact and disease progression loops and updates the agents state at the next time step. It also updates all other states as follows.
  a. *Individuals in* **A**_S_ *at time t*. For the *N*_S_(*t*) individuals in **A**_S_ at time *t*, we have 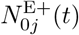 enter set 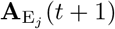 and we update

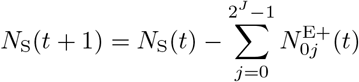

where Eq. A.20 ensures that *N*_S_(*t* + 1) ≥ 0
  b. *Individuals in* **A**_S_ *that are infected again over* [*t, t* + 1). These individuals can become reinfected as calculated in the contact loop. Those that become reinfected with variant *j, j* = 0, …, 2^*J*^ − 1 enter state **E**_*j*_(*t* + 1, *t* + 1) at time *t* + 1.
  c. *Updating the immunity of individuals in* **A**_S_. Every individual within **A**_S_ at time *t* must have its immunity status updated so that for *j* = 0, …, 2^*J*^ − 1, if A_*i*_ is in state V_*j*_(*t, τ*_*ij*_) at time *t* then it transfers to state V_*j*_(*t* + 1, *τ*_*ij*_) at time *t* + 1, even if reinfected, as in b.) above.
  d. *Transfer from* **S** *to* **A**. The 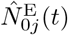 computed in Eq. A.21 become newly listed members of the set **A** by entering state **E**_*j*_(*t* + 1, *t* + 1), *j* = 0, …, 2^*J*^ − 1. This involves updating the equations for *N*_S_(*t*) and *N*_A_(*t*), including taking account of the number of individuals Δ*N*_D_(*t*) that died from the disease in the immediate time period, i.e.:

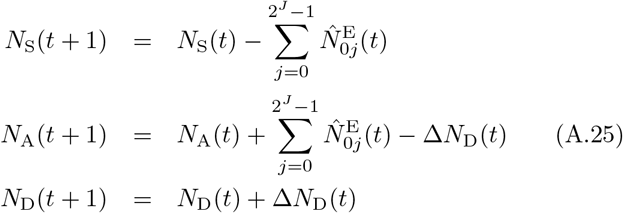
  e. Along with input parameter values *t*_vac___on_ ≥ 0, *t*_vac___off_ and *p*_*v*_ ∈ − [0, 0.1], we also need to specify the valency of the vaccination by selecting 1 to 4 numbers that take on values 0, …, 2^*J*^ − 1 (if more valencies are needed than 4, then the platform needs to be modified accordingly). We also need specify whether *N*_select_ will just be individuals in the set **S**(*t*) (*N*_select_ = *N*_S_) or will be any individual other than those in the set **A**_I_(*t*) (*N*_select_ = *N*_S_ + *N*_A_ − *N*_I_).

In Algorithm 1 we summarise the steps of the simulation algorithm, as described in this section. On the right we report the name and numbering of the subsections while in the for loops we list the various steps respecting the item letters. Note that technical steps not explicitly described in the text (e.g. store updates, store set progression) do not present letters or numbers. The time set is defined with *T* while to describe temporal progression of set **S, A** and **D** we use the symbols 𝒮, 𝒜, 𝒟 respectively.

#### A.4 Estimation of *R*_0_

In a finite population, a pathogen can emerge from a single infection with probability *p*_outbreak_ = 1 − 1*/R*_0_ if *R*_0_ > 1, otherwise an outbreak will not occur [42]. Thus we can estimate *R*_0_ if we have an estimate of *p*_outbreak_ and the use the following relationship to compute *R*_0_

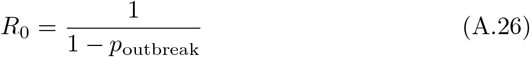

For the set of parameters listed in Table 2, from 100 runs (runtime seed goes from 0 to 99 in 100 separate simulations) of the single strain case, we estimated *p*_outbreak_ ≈ 0.68 from the the proportion of simulations that had positive prevalence after 100 days. From Eq. A.26 this implies that *R*_0_ ≈ 3.1, with a 95% confidence interval of *R*_0_ ∈ [2.4, 4.3] (this of course can be narrowed down with additional simulations, but a more precise estimate is not particularly relevant to our illustrative results).

##### Algorithm 1: Summary of simulation algorithm

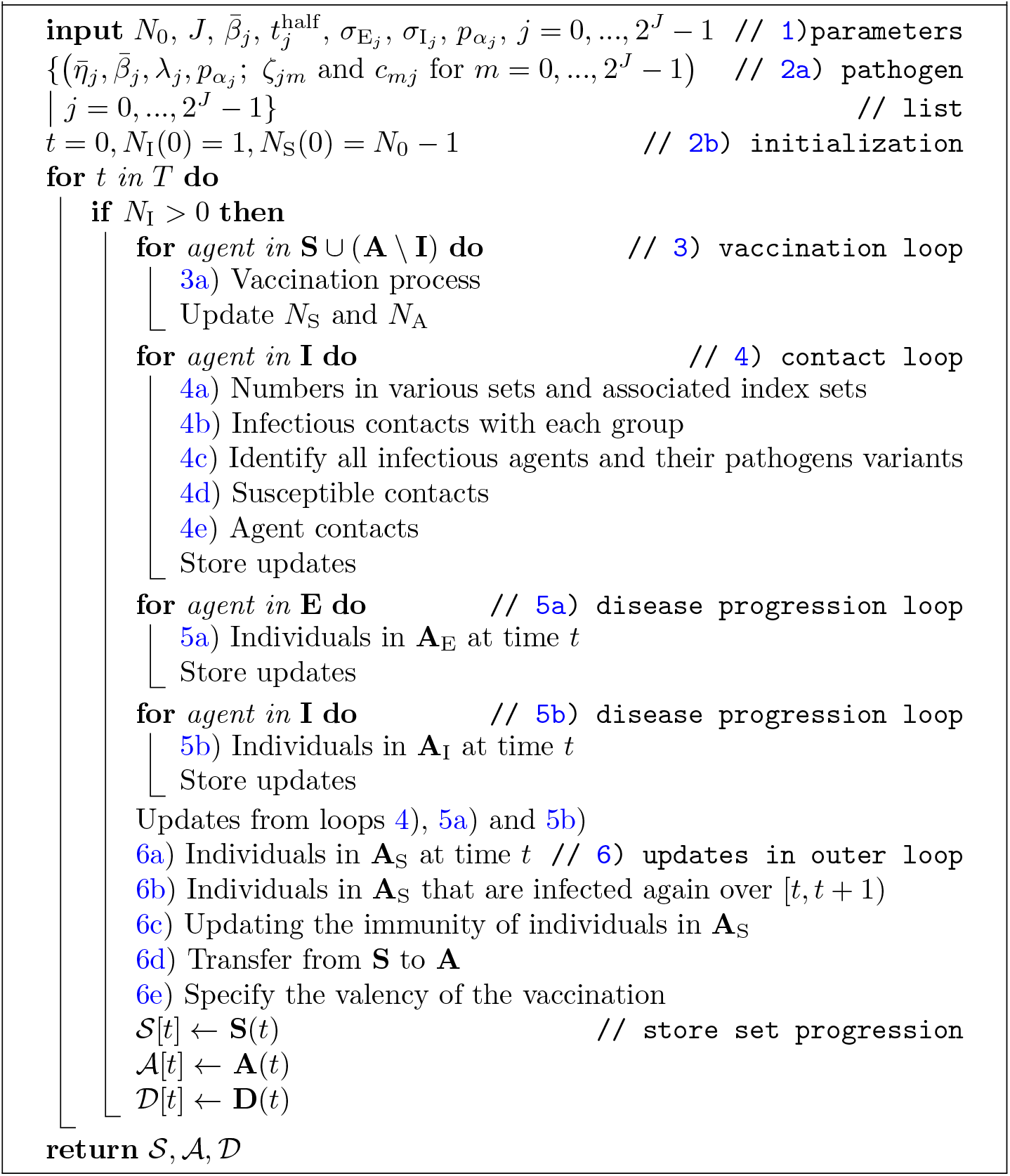

### B RAMP Details

#### B.1 General description

An open and expressive design of the model platform aids in and encourages exploration and experimentation. The RAMP design augments a desktop simulation platform with several novel features that increase flexibility and expressiveness, and promote experimentation and interoperability with other platforms. These include an API (“application programming interface”) fully supporting remote operation and direct retrieval of data for external processing on other platforms, such as Python, Javascript or the R statistical platform. The API can also be accessed by an onboard scripting interface that uses the Nashorn Javascript engine.

Additionally, using a novel design, elements of the internal algorithm are exposed for possible reprogramming in a secure fashion that will not damage the overall system. These runtime alternative modules (RAMs) may also be controlled from the API to facilitate selective algorithm redefinition during the run of the simulation.

Use of the RAMP features require some experience with scripting and/or Java coding, however the resulting modifications to the algorithm can be of great significance. The RAM platform is implemented to support program redefinition with no risk to damaging the underlying code base. It should be accessible to anyone with moderate scripting experience.

A major goal of the RAMP project is to prepackage these functionalities so that they can be readily deployed as part of simulation system design. This goal has been partially realized with respect to the RAM platform: annotations can be added to the simulator’s source code that direct the automatic generation of Java code to integrate into the simulations’ source and provide the functionality.

The following discussion assumes some familiarity with script or program development.

#### B.2 Runtime alternative modules

Figure B.1 shows the RAM redefinition frame. The available RAMs appear as radio buttons along the bottom of the frame. Each RAM is a set of options for defining a relatively short Java method implementing some key aspect of the simulation algorithm. For example, included in this simulation are the implementation for cross immunity given in Eq. 1; the implementation for *β* given in Eq. A.11; and the implementation for *ϕ* given in Eq. A.8; etc. Each RAM initially contains only a single option, Option 0, the default, internally defined implementation. Option 0 cannot be edited and appears for reference purposes only.

Additional options may be added to each RAM containing code redefining the method. Two editor panes and one console pane are stacked in the frame and display the code and output of the RAM. These panes show the content associated with the currently selected RAM and option. The top editor pane contains the code for the method being redefined. The second editor pane contains definitions of any new help functions required by the definition in the top pane. The console pane contains messages and output that are useful during the development of the option. For convenience, a “Load Default” button initializes the editor to an editable version of the Option 0 default to use as a starting point.

**Figure B.1:**
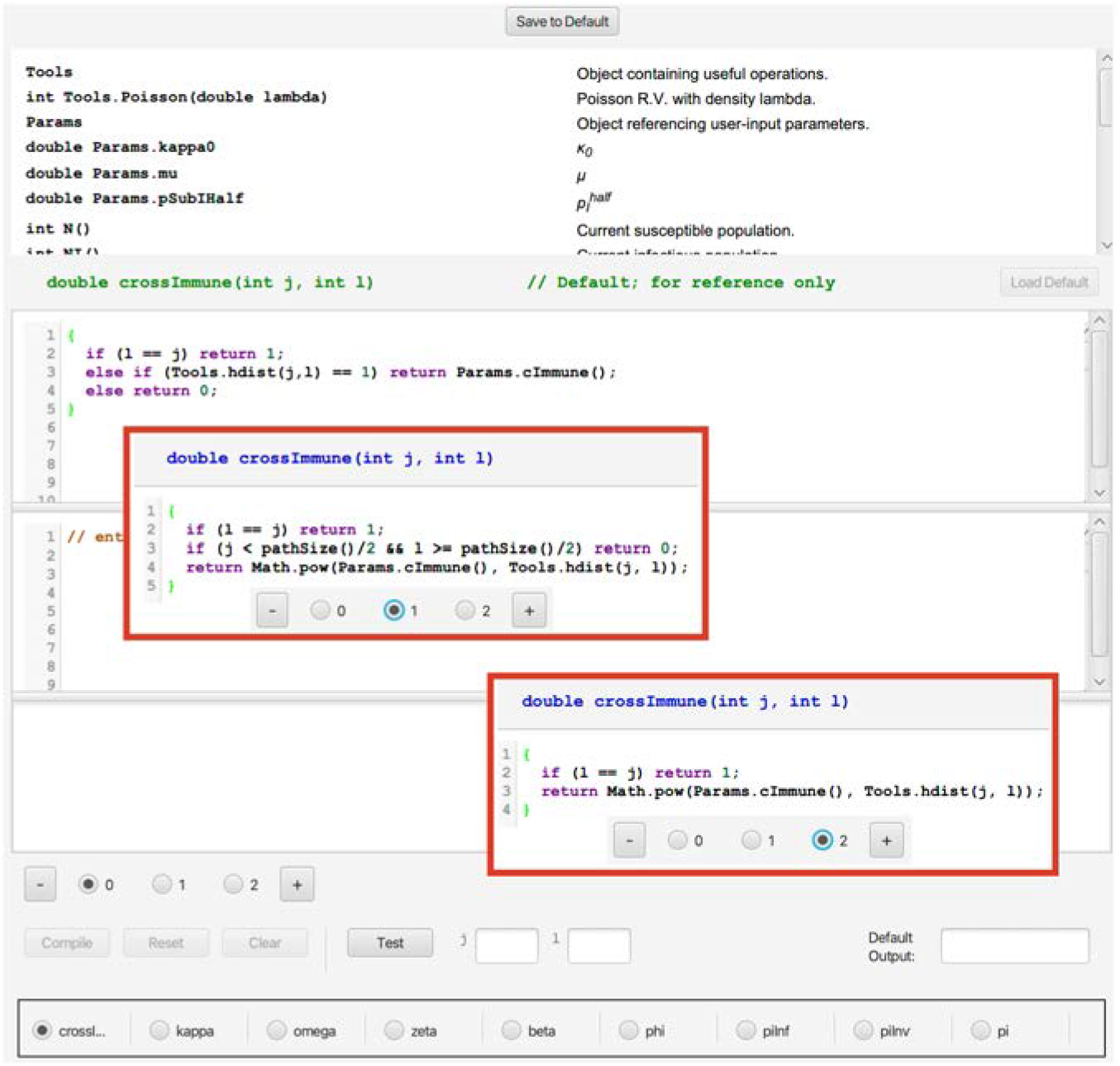
RAM frame shows the implementation of a generic nearest-neighbor cross-immunity formulation **C** as the default. In the red-bordered insets are the cascading cross-immunity without (blue highlighted radio button 1) and with an idealized escape mutation (blue highlighted radio button 2), as formulated in Eqs. 1 and 2 respectively. Note the “+” button which allows for an unlimited number of alternatives to be set up using consecutive integer numbers for the new radio-buttons that appear and pertain to the selection of each alternative. Also, note the bottom list of functions that can be altered at runtime. The “load default” button on the upper left-hand side allows the user, when starting a new alternative, to insert the default code (which is immutable in radio-button 0) as a starting point. The frame also documents a list of terms in the upper panel that can be used to build any function.

Figure B.1 shows the Option 0 default definition for the cross immunity matrix function, as described by Eq. 1. Clicking the “+” button produced two new options, which appear in the insets. These option implement the alternative cascading cross immunity schemes presented in Eqs. 1 and 2. code appears below:

~~~
double crossImmune(int j, int k) {
    if (l == j) return 1;
    if (j < pathSize()/2 && l >= pathSize()/2) return 0;
    return Math.pow(Params.cImmune(), Tools.hdist(j, l));
}
~~~

Note that we have substituted the function `pathSize` for a hard-coded value of 63. `pathSize` returns the number of pathogens, allowing us to use this formulation for any choice of entropy. Documentation for `pathSize` is at the top of the window in the list of available help functions and parameters. There is also more extensive documentation in a separate user guide (see Fig. B.2).

**Figure B.2:**
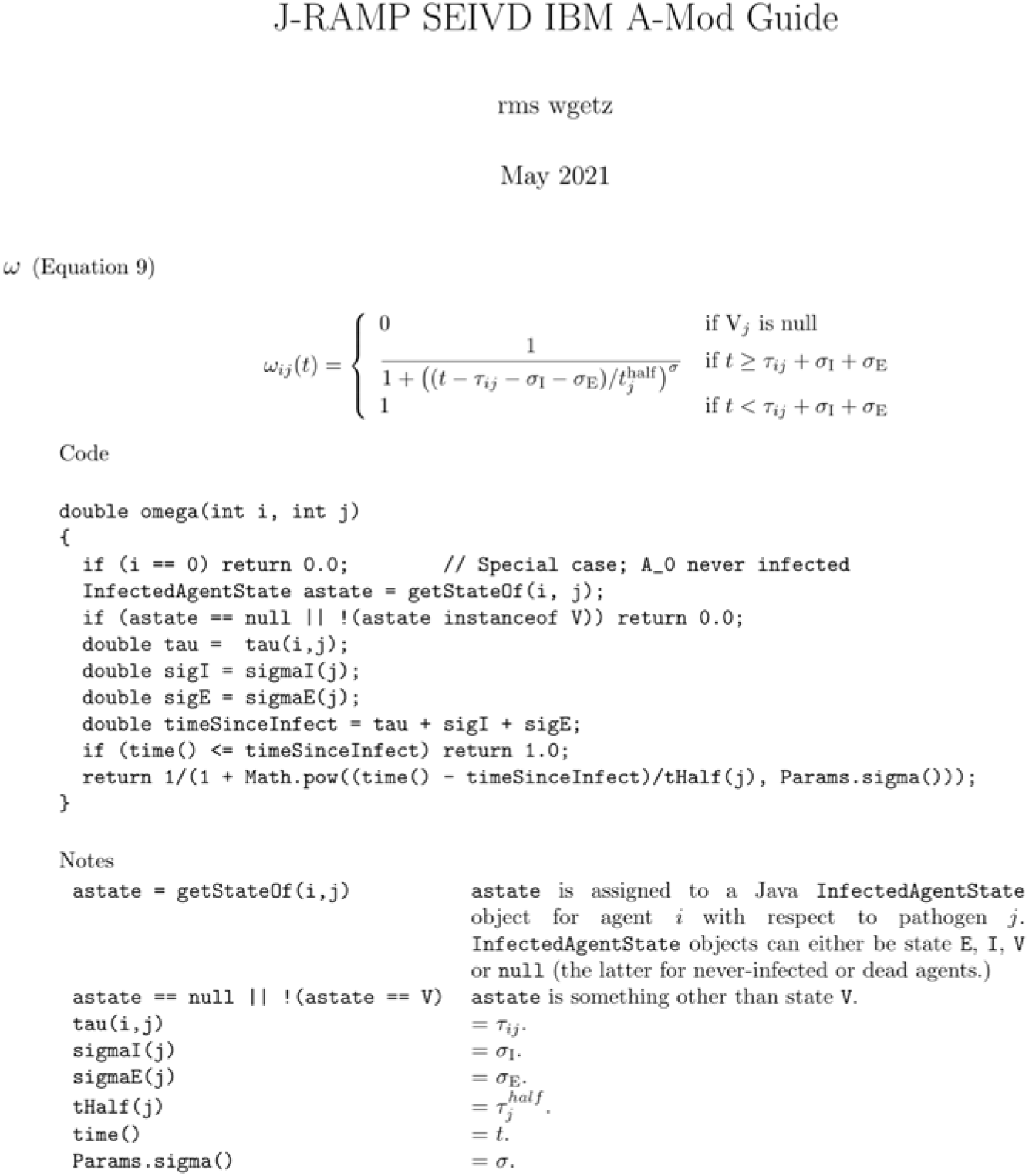
A description from our User Guide of the immunity waning function *ω*.

The platform duplicates a mini-development environment for building alternative definitions. Once code has been entered the “Compile” button checks the legality of the code and makes it available for use at runtime. Legally compiled code will produce a “Compilation Successful” message. Errors will appear with line numbers if they occur. Once the code is legal, the “Test” button can be used with actual parameters entered into the small text fields to determine correctness of the code. It is also possible to include `print` and `println` statements in the code during development to further check correctness. Output from print statements will appear in the bottom console window. The entire RAM set can be saved and will reappear during subsequent launches of the simulator platform.

To use an alternate RAM definition at runtime simply select the dM-SEIRed option. (Selected options will be restored from a saved RAM set during subsequent launches.) The system will compile any uncompiled code the first time it is accessed. If an error occurs during a runtime compilation an alert will notify the user that the system is returning to the default definition of that RAM. At no time is the internal logic of the program overridden.

Finally, RAM option selection is part of the API described in the next section. This means that a script may run a simulation selecting different options at different points in time, using logic that considers the state of the model. For example, such an adaptive protocol might be appropriate for determining the contact rate *κ*.

#### B.3 Application programming interface

The API is a simple bytecode^1^ called BPL (Blackbox Programming Language) that addresses all available user interactions with the simulator. Instructions fall into three categories: parameter assignment and retrieval; simulator operation; and data retrieval. A complete list of instructions is shown in Fig. B.3. Instructions are comprised of opcodes (e.g., `reset, step, get`) followed by 0 or more arguments. Every BPL operation returns a result, even if empty, for synchronization purposes. A string consisting of a sequence of opcodes and arguments may be submitted to the BPL interpreter, an example of which is shown in the notes in Fig. B.3.

##### Parameter assignment and retrieval

Every user-configurable element (including random number generator seeds) is addressed from BPL using a unique three-letter “airport code” (see Table. B.31). Additionally, pathogens are addressed by their id number (0 to 2^*J*^ − 1) and agent states using identifiers S, E, I, V, DI+ and DD (the latter two represent ΔI and ΔD, respectively). RAM options are addressed in `setOption` and `getOption` using the name of the RAM (e.g., “crossImmune”). Get and set operations can be used on each of these with the exception of ENT (variant entropy), which is read-only.

##### Simulator operation

Simulation runs begin by executing the BPL `reset` instruction, followed by **step, run for** or **run**. The BPL interpreter operates synchronously with the simulator by waiting to process subsequent commands during a simulation run. Operational instructions can be interspersed with parameter set/get or data retrieval to use in runtime decision-making. Note that the **reset** operation restores the simulator to its state at the time of the last **reset**, so that no parameter changes made during a run are persistent.

##### Data retrieval

Operations to obtain the current population in each state, and to retrieve the runtime population history of each state and pathogen are also included. These can be easily transformed into R data frames, for example, for further analysis.

Scripting can be deployed using either one of the two on-board script interpreter interfaces, or remotely from another platform using drivers provided with the simulator. The remote drivers use TCP/IP *sockets*. Sockets are integral to Internet communication, and so are found on any system supporting the Internet. In this case the simulator acts as a server fielding API requests from the remote drivers.

On our main dashboard, we provide two scripting windows that are opened using the “S On” and “JS On” buttons (see button second and third from left at bottom of Fig. 2A). The former allows the user to write simulation driver scripts directly as command strings. (The commands listed in Fig. B.3 are accessed by pressing the “Command Reference” button in the “S On” window.) This window is used primarily to test and monitor scripts intended to be deployed on a remote platform. The JS window contains a Nashorn Javascript interpreter enhanced to accept and execute BPL operations. Scripts can developed, saved, and used to drive the simulator from this interpreter. For example, Fig. B.4, lists the code used to implement the adaptive vaccination programs. The `SEIV` object referenced in this code contains methods corresponding to the BPL operations detailed in Fig. B.3.

**Table B1:**
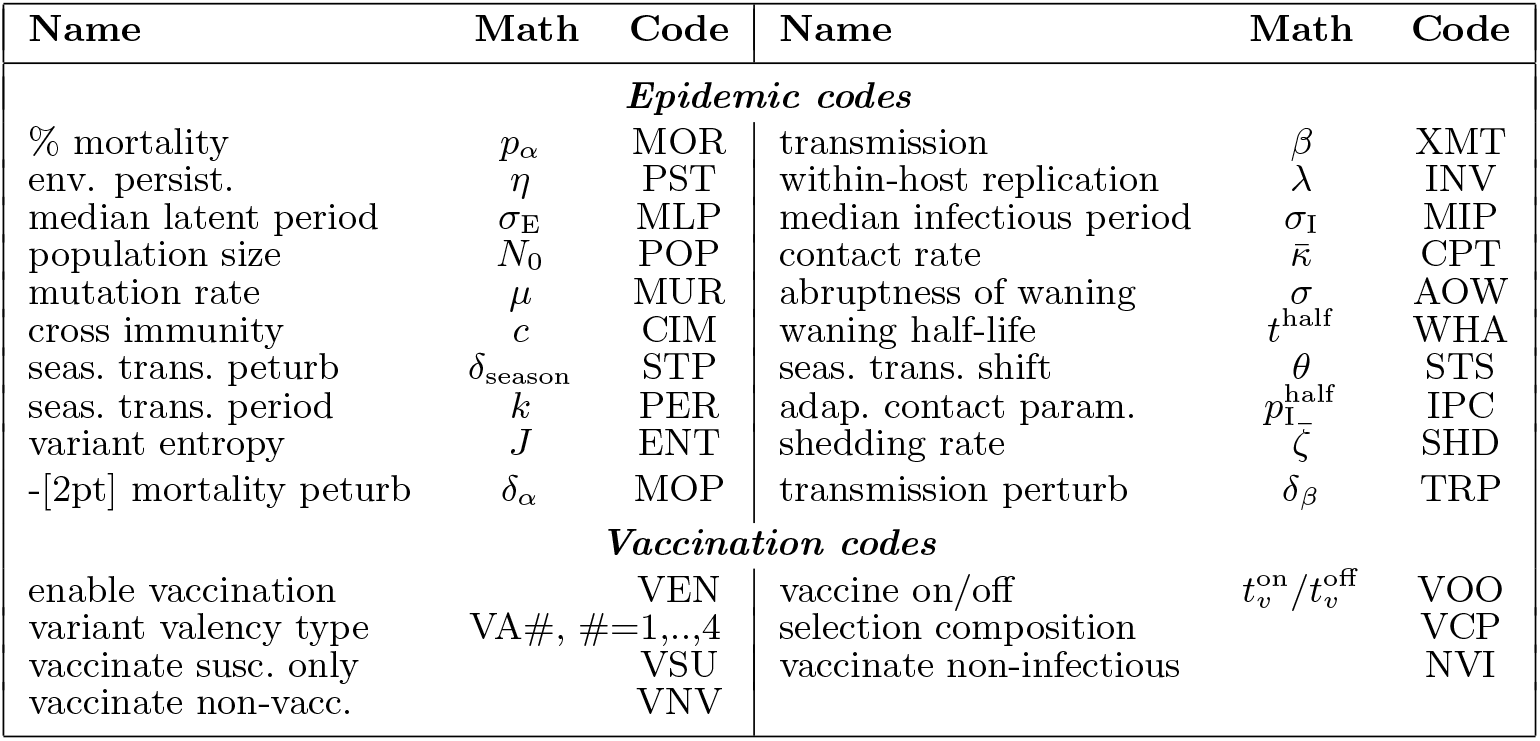
Airport codes for parameter and variables used in the model algorithm

**Figure B.3:**
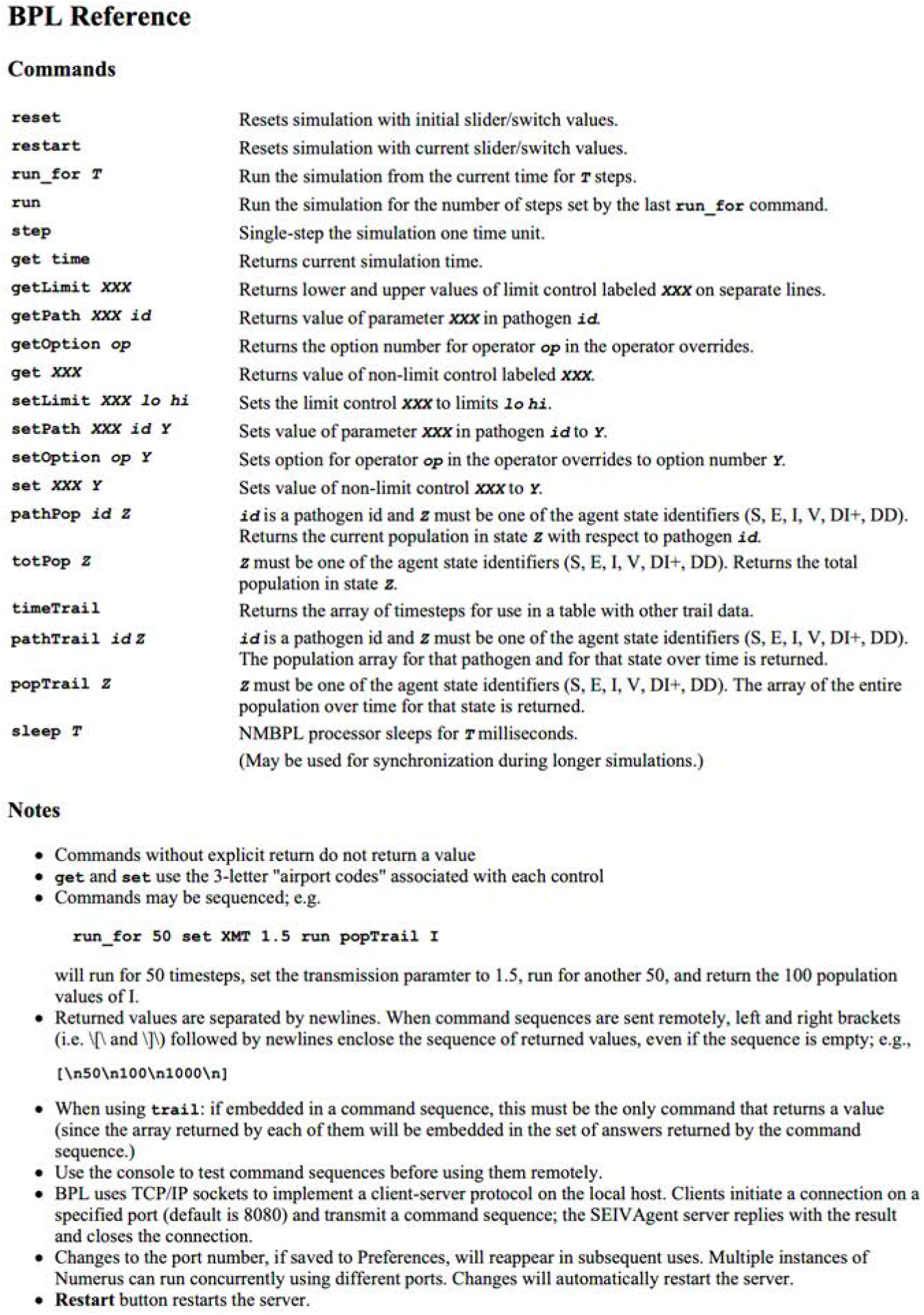
The list of Blackbox Programming Language (BPL) commands that can be used to write a simulation driver script, using the three-letter “airport codes” listed in Table A1 to access the parameters and variables in our coded algorithm. This list of commands can be accessed using the “Command Reference” button at the bottom of the Scripting Window.

**Figure B.4:**
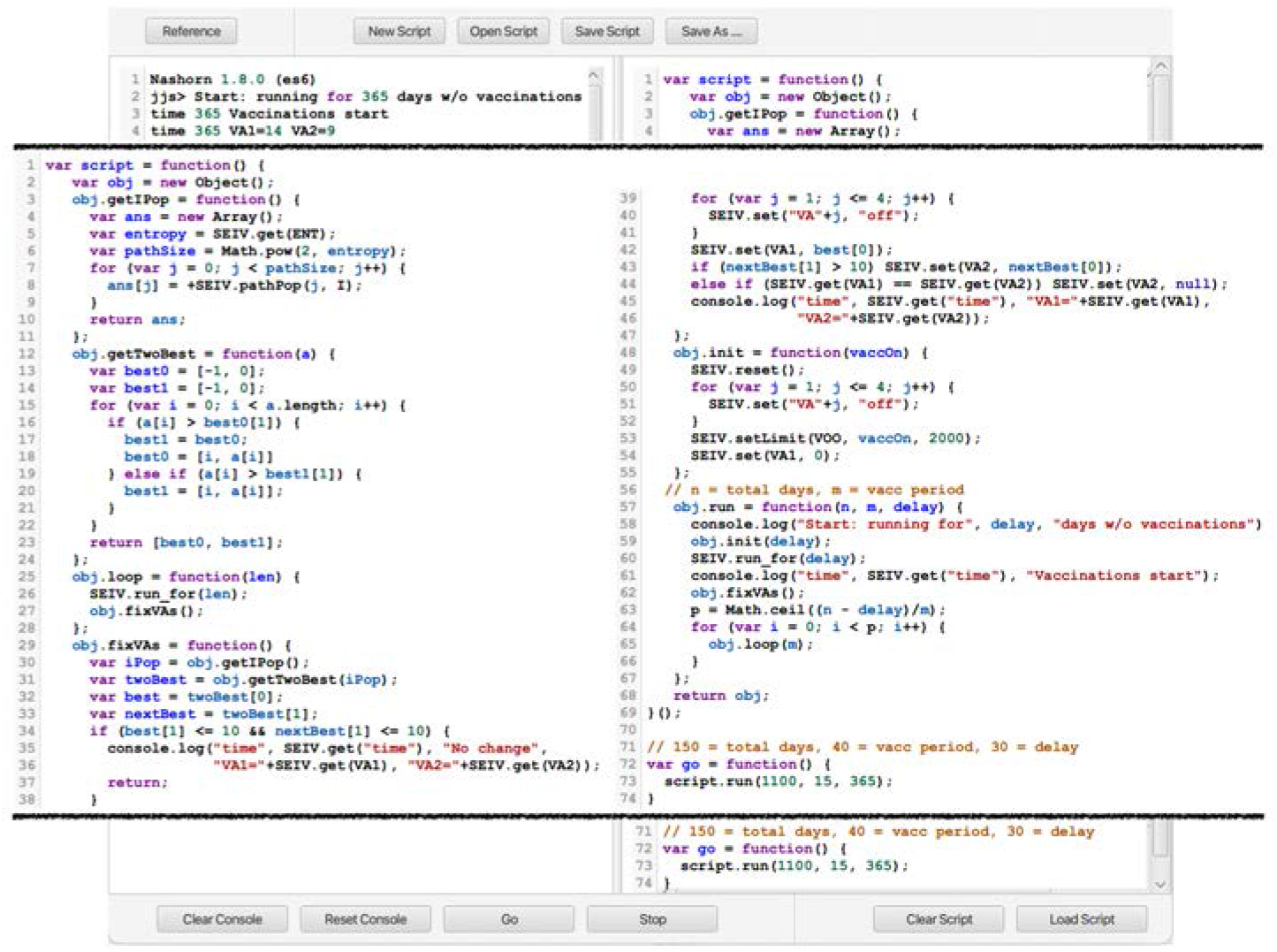
The JS scripting window accessed by selecting the “JS On” button in the main dashboard (see button second from left at bottom of Fig. 2A). The script shown here (broken into two columns 1-74 and overlaid over JS window) was used to execute the adaptive vaccination strategy discussed in the main text.

#### B.4 R Integration

As previously mentioned, the API supports remote control of the simulator from independent platforms using the operating system’s socket interface^2^. Of partic-ular interest is integration with the R statistical programming environment. An R-package called “seiv” acts as a driver by synchronously issuing BPL command strings and waiting for results. Consequently, a simulation can be driven entirely from within the R platform, treating the simulator as a “virtual package”.

Fig. B.5 shows the code used to run the simulator multiple times with different random number generator seeds. Following each run, the time history of the population in the I, DI+ and DD states is extracted directly to an R data frame (without the need to save, for example, in a comma-separated list). At the end of the run sequence the data frame is used to build the plots shown in

Fig. 3D & E. R could be used in a more direct way by analyzing data at various points throughout a single run and adjusting parameters programmatically, similar to the adaptive vaccination strategy carried out in Javascript, only taking advantage of the R environment’s powerful toolkit.

**Figure B.5:**
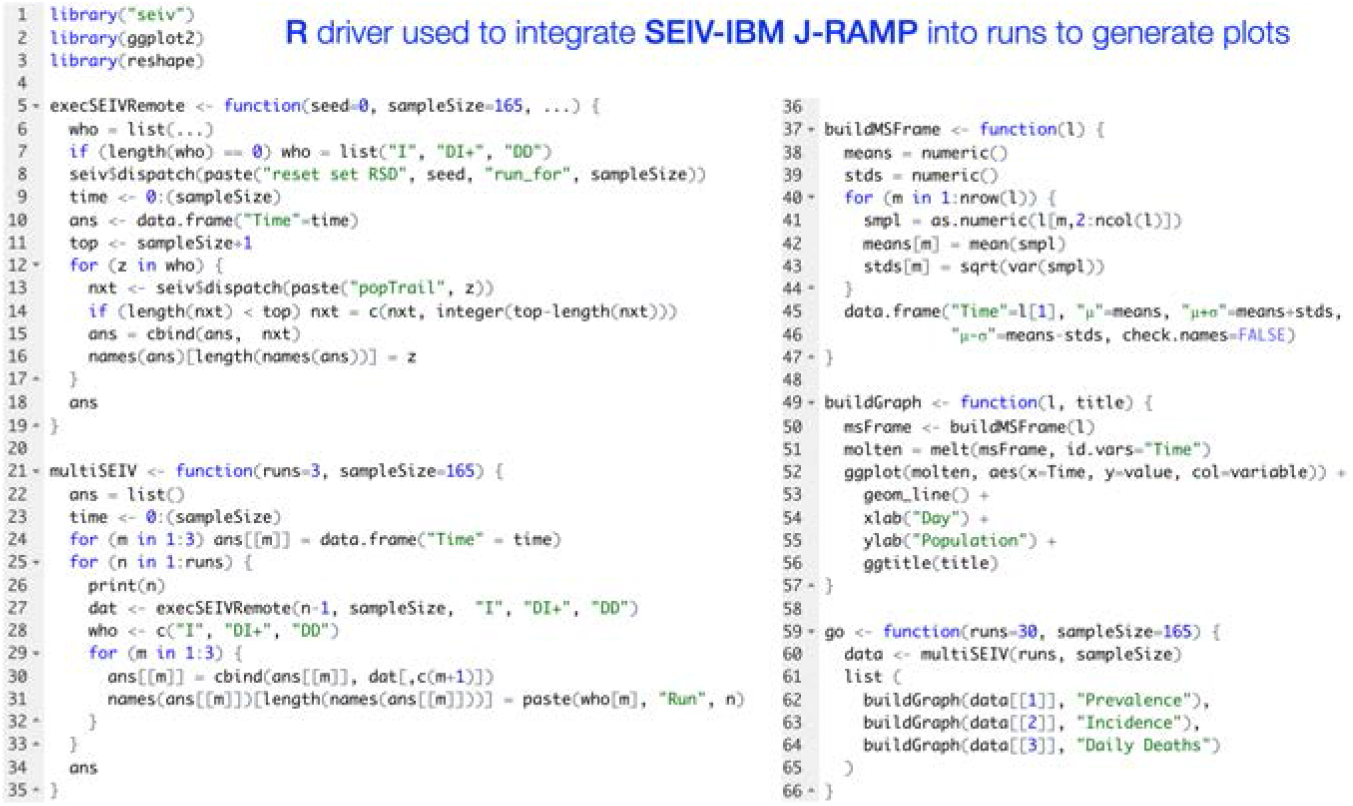
Our M-SEIR can be treated as an R-package called “seiv” and run as such in conjunction with other packages, such as ggplot2 and reshape to conduct multiple simulations and then carry out data and statistical analyses of the simulation results. The code shown here was used to produce the plots illustrated in Fig. 3D & E.

## Footnotes

1 a *bytecode* is computer source code that is processed immediately by a program, usually referred to as an interpreter or virtual machine.

2 By using internet sockets, the simulator and R platform could conceivably run on different systems.

## Notes

### Competing Interest Statement

The authors have declared no competing interest.

### Summary of Updates

We have changed the title, abstract and focus on the paper. We have run a new set of scenarios relevant to the new focus of the publication.

## References

[1] Kermack WO, McKendrick AG. A contribution to the mathematical theory of epidemics. Proceedings of the royal society of london Series A, Containing papers of a mathematical and physical character. 1927;115(772):700–721.

[2] Hethcote HW. The mathematics of infectious diseases. SIAM Review. 2000;42(4):599–653.

[3] Koelle K, Cobey S, Grenfell B, Pascual M. Epochal evolution shapes the phylodynamics of interpandemic influenza A (H3N2) in humans. Science. 2006;314(5807):1898–1903.

[4] Gilchrist MA, Sasaki A. Modeling host–parasite coevolution: a nested approach based on mechanistic models. Journal of Theoretical Biology. 2002;218(3):289–308.

[5] Balcan D, Goncalves B, Hu H, Ramasco JJ, Colizza V, Vespignani A. Modeling the spatial spread of infectious diseases: The GLobal Epidemic and Mobility computational model. Journal of computational science. 2010;1(3):132–145.

[6] Keeling MJ. The effects of local spatial structure on epidemiological invasions. Proceedings of the Royal Society of London B: Biological Sciences. 1999;266(1421):859–867.

[7] Keeling MJ, Eames KT. Networks and epidemic models. Journal of the Royal Society Interface. 2005;2(4):295–307.

[8] Bauer AL, Beauchemin CA, Perelson AS. Agent-based modeling of host– pathogen systems: The successes and challenges. Information sciences. 2009;179(10):1379–1389.

[9] Cuevas E. An agent-based model to evaluate the COVID-19 transmission risks in facilities. Computers in biology and medicine. 2020;121:103827.

[10] Rambaut A, Holmes EC, O’Toole Á, Hill V, McCrone JT, Ruis C, et al. A dynamic nomenclature proposal for SARS-CoV-2 lineages to assist genomic epidemiology. Nature microbiology. 2020;5(11):1403–1407.

[11] Koopman JS, Simon CP, Getz WM, Salter R. Modeling the population effects of escape mutations in SARS-CoV-2 to guide vaccination strategies. Epidemics. 2021;p. 100484.

[12] Aslaner H, Aslaner HA, Gökçek MB, Benli AR, Yıldız O. The effect of chronic diseases, age and gender on morbidity and mortality of covid-19 infection. Iranian Journal of Public Health. 2021;50(4):721.

[13] Getz WM, Salter R, Mgbara W. Adequacy of SEIR models when epidemics have spatial structure: Ebola in Sierra Leone. Philosophical Transactions of the Royal Society B. 2019;374(1775):20180282.

[14] Larsen LG, Eppinga MB, Passalacqua P, Getz WM, Rose KA, Liang Appropriate complexity landscape modeling. Earth-science reviews. 2016;160:111–130.

[15] Getz WM, Salter R, Luisa Vissat L, Horvitz N. A versatile web app for identifying the drivers of COVID-19 epidemics. Journal of Translational Medicine. 2021;19(1):1–20.

[16] Rahimi I, Chen F, Gandomi AH. A review on COVID-19 forecasting models. Neural Computing and Applications. 2021;p. 1–11.

[17] Chretien JP, George D, Shaman J, Chitale RA, McKenzie FE. Influenza forecasting in human populations: a scoping review. PloS one. 2014;9(4):e94130.

[18] Getz WM, Gonzalez JP, Salter R, Bangura J, Carlson C, Coomber M, et al. Tactics and strategies for managing Ebola outbreaks and the salience of immunization. Computational and mathematical methods in medicine. 2015;2015.

[19] Chen PZ, Bobrovitz N, Premji Z, Koopmans M, Fisman DN, Gu FX. Heterogeneity in transmissibility and shedding SARS-CoV-2 via droplets and aerosols. eLife. 2021 apr;10:e65774. Available from: https://doi.org/10.7554/eLife.65774.

[20] van Doremalen N, Bushmaker T, Morris DH, Holbrook MG, Gamble A, Williamson BN, et al. Aerosol and Surface Stability of SARS-CoV-2 as Compared with SARS-CoV-1. New England Journal of Medicine. 2020;382(16):1564–1567. Available from: https://doi.org/10.1056/NEJMc2004973.

[21] Maginnis MS. Virus–receptor interactions: the key to cellular invasion. Journal of molecular biology. 2018;430(17):2590–2611.

[22] Challen R, Brooks-Pollock E, Read JM, Dyson L, Tsaneva-Atanasova K, Danon L. Risk of mortality in patients infected with SARS-CoV-2 variant of concern 202012/1: matched cohort study. bmj. 2021;372.

[23] Sariol A, Perlman S. Lessons for COVID-19 immunity from other coronavirus infections. Immunity. 2020;.

[24] Eguia RT, Crawford KH, Stevens-Ayers T, Kelnhofer-Millevolte L, Greninger AL, Englund JA, et al. A human coronavirus evolves antigenically to escape antibody immunity. PLoS pathogens. 2021;17(4):e1009453.

[25] Alizon S, Luciani F, Regoes RR. Epidemiological and clinical consequences of within-host evolution. Trends in microbiology. 2011;19(1):24–32.

[26] Fenichel EP, Castillo-Chavez C, Ceddia MG, Chowell G, Parra PAG, Hickling GJ, et al. Adaptive human behavior in epidemiological models. Proceedings of the National Academy of Sciences. 2011;108(15):6306–6311.

[27] Gozzi N, Bajardi P, Perra N. The importance of non-pharmaceutical interventions during the COVID-19 vaccine rollout. medRxiv. 2021;Available from: https://www.medrxiv.org/content/early/2021/01/09/2021.01.09.21249480.

[28] Kumar A, Dowling WE, Román RG, Chaudhari A, Gurry C, Le TT, et al. Status Report on COVID-19 Vaccines Development. Current Infectious Disease Reports. 2021;23(6):1–12.

[29] Jefferies JM, Clarke SC, Webb JS, Kraaijeveld AR. Risk of red queen dynamics in pneumococcal vaccine strategy. Trends in microbiology. 2011;19(8):377–381.

[30] Willem L, Verelst F, Bilcke J, Hens N, Beutels P. Lessons from a decade of individual-based models for infectious disease transmission: a systematic review (2006-2015). BMC infectious diseases. 2017;17(1):612.

[31] Chowell G, Sattenspiel L, Bansal S, Viboud C. Mathematical models to characterize early epidemic growth: A review. Physics of life reviews. 2016;18:66–97.

[32] Lavine JS, Bjornstad ON, Antia R. Immunological characteristics govern the transition of COVID-19 to endemicity. Science. 2021;371(6530):741–745.

[33] Koyama T, Weeraratne D, Snowdon JL, Parida L. Emergence of drift variants that may affect COVID-19 vaccine development and antibody treatment. Pathogens. 2020;9(5):324.

[34] Getz WM, Lloyd-Smith JO. Basic methods for modeling the invasion and spread of contagious diseases. DIMACS Series in Discrete Mathematics and Theoretical Computer Science. 2006;71:87.

[35] McCallum H, Barlow N, Hone J. How should pathogen transmission be modelled? Trends in ecology &amp; evolution. 2001;16(6):295–300.

[36] Anderson RM, May RM, et al. Coevolution of hosts and parasites. Parasitology. 1982;85(Pt 2):411–426.

[37] Getz WM, Salter R, Muellerklein O, Yoon HS, Tallam K. Modeling epidemics: A primer and Numerus Model Builder implementation. Epidemics. 2018;25:9–19.

[38] Getz WM, Salter R, Lyons AJ, Sippl-Swezey N. Panmictic and clonal evolution on a single patchy resource produces polymorphic foraging guilds. PloS one. 2015;10(8):e0133732.

[39] Hussein M, Toraih E, Elshazli R, Fawzy M, Houghton A, Tatum D, et al. Meta-analysis on serial intervals and reproductive rates for SARS-CoV-2. Annals of surgery. 2021;273(3):416–423.

[40] Lau H, Khosrawipour T, Kocbach P, Ichii H, Bania J, Khosrawipour V. Evaluating the massive underreporting and undertesting of COVID-19 cases in multiple global epicenters. Pulmonology. 2021;27(2):110–115.

[41] Karlinsky A, Kobak D. Tracking excess mortality across countries during the COVID-19 pandemic with the World Mortality Dataset. eLife. 2021;10:e69336.

[42] Hartfield M, Alizon S. Introducing the outbreak threshold in epidemiology. PLoS pathogens. 2013;9(6):e1003277.

[43] Getz WM, Luisa Vissat L, Salter R. A Contact-Explicit Covid-19 Epidemic and Response Assessment Model. medRxiv. 2020;.

[44] Greaney AJ, Starr TN, Gilchuk P, Zost SJ, Binshtein E, Loes AN, et al. Complete mapping of mutations to the SARS-CoV-2 spike receptorbinding domain that escape antibody recognition. Cell host & microbe. 2021;29(1):44–57.

[45] Funk S, King AA. Choices and trade-offs in inference with infectious disease models. Epidemics. 2020;30:100383.

[46] Suthar MS, Zimmerman MG, Kauffman RC, Mantus G, Linderman SL, Hudson WH, et al. Rapid generation of neutralizing antibody responses in COVID-19 patients. Cell Reports Medicine. 2020;1(3):100040.

[47] Muruato AE, Fontes-Garfias CR, Ren P, Garcia-Blanco MA, Menachery VD, Xie X, et al. A high-throughput neutralizing antibody assay for COVID-19 diagnosis and vaccine evaluation. Nature communications. 2020;11(1):1–6.

[48] Jeyanathan M, Afkhami S, Smaill F, Miller MS, Lichty BD, Xing Z. Immunological considerations for COVID-19 vaccine strategies. Nature Reviews Immunology. 2020;20(10):615–632.

[49] Tkachenko AV, Maslov S, Elbanna A, Wong GN, Weiner ZJ, Goldenfeld Time-dependent heterogeneity leads to transient suppression of the COVID-19 epidemic, not herd immunity. Proceedings of the National Academy of Sciences. 2021;118(17).

[50] Anderson RM. The impact of vaccination on the epidemiology of infectious diseases. In: The Vaccine Book. Elsevier; 2016. p. 3–31.

[51] Kramer A, Schwebke I, Kampf G. How long do nosocomial pathogens persist on inanimate surfaces? A systematic review. BMC infectious diseases. 2006;6(1):1–8.

[52] Liu X, Huang J, Li C, Zhao Y, Wang D, Huang Z, et al. The role of seasonality in the spread of COVID-19 pandemic. Environmental research. 2021;195:110874.

[53] Valtonen A, Molleman F, Chapman CA, Carey JR, Ayres MP, Roininen H. Tropical phenology: Bi-annual rhythms and interannual variation in an Afrotropical butterfly assemblage. Ecosphere. 2013;4(3):1–28.

[54] Lythgoe KA, Hall M, Ferretti L, de Cesare M, MacIntyre-Cockett G, Trebes A, et al. SARS-CoV-2 within-host diversity and transmission. Science. 2021;372(6539). Available from: https://science.sciencemag.org/content/372/6539/eabg0821.

